# The role of socioeconomic deprivation on inequities in care outcomes of haematological malignancies, treated with stem cell transplant in the UK: A systematic Review Protocol

**DOI:** 10.1101/2025.09.19.25336176

**Authors:** Zareen Deplano, Samuel Cusworth, Carolyn Doree, James Griffin, Nicola J Adderley, Joht Singh Chandan

## Abstract

**Introduction:** Stem cell transplant (SCT) is a potential cure for haematological malignancies whose outcomes are predicted to be poor on chemotherapy alone and cases difficult to treat using standard of care treatment. There is growing evidence to show that not all patients who would benefit from SCT, receive it due to inequities in the treatment pathway. The role of socioeconomic deprivation on inequities in the SCT setting is unclear due to conflicting findings. We aim to synthesis the UK evidence base in this area to guide further research and policy to improve survival for underserved patients.

**Method and analysis:** Primary electronic searches will be performed in bibliographic databases including MEDLINE, Embase, Proquest Databases, Central & CDSR, Stem Cell Evidence and Web of Science and Google Scholar for grey literature. Eligibility criteria will include observational studies of UK patients diagnosed with haematological malignancies, treated with SCT, where outcomes are reported according to socioeconomic deprivation. Two independent reviewers will screen each study and assess the quality of all included studies be using the Newcastle-Ottawa grading system. Outcomes of interest include delay in diagnosis/referral, treatments/interventions received, referral to transplant, presence of co-morbidities affecting transplant eligibility, access to transplant, transplant utilisation rates, waiting time to transplant and survival rates. Inequities in SCT due to socioeconomic deprivation will be narratively summarised. Meta-analysis will be conducted dependent on data availability. This protocol has been registered with PROSPERO (2024: CRD42024620592).

**Ethics and dissemination:** Ethical approval is not required for this study as it is a secondary analysis of published data. The findings will be submitted for peer-reviewed publication, shared at conference presentations, and disseminated to both clinical and patient groups including National (Black, Asian, Mixed Race, and Minority Ethnic) BAME Transplant Alliance (NBTA) and the National Institute for Health Research (NIHR) Blood and Transplant Research Unit (BTRU) in Precision Transplant and Cellular Therapeutics.

**Article Summary:** *Strengths and limitations of this study:*

- This review will provide the first comprehensive and systematic summary of inequities due to socioeconomic deprivation in the stem cell transplant setting in the UK, following the Preferred Reporting Items for Systematic Reviews and Meta Analyses (PRISMA) guidelines
- This review will focus on data collected from studies performed in a UK population only. The findings can be applied to a UK population but application to other countries will be limited.
- Patients and the public participated in the development of the review and will support interpretation of findings and dissemination to the wider public.

## Introduction

Haematological malignancies are cancers of the blood (leukaemia), lymph nodes (lymphomas - Hodgkin and non-Hodgkin) or bone (myelomas) (1,2). They represent the fifth most common cancer type and the third largest cause of cancer-related death, with 41,000 diagnoses and 16,000 deaths yearly in the UK (3–5). Stem cell transplantation (SCT) is a potentially curative treatment for those patients with poor prognosis on chemotherapy alone and cases difficult to treat using standard of care treatment (6–8). The most common clinical indications for SCT are acute Myeloid Leukaemia (AML), acute Lymphoid Leukaemia (ALL) and myelodysplastic Syndrome (MDS) and Myeloma In adults with AML in first complete remission (CR1), SCT has been shown to reduce the risk of disease relapse by 60% compared to intensive chemotherapy alone A systematic review and meta-analysis by Koreth et al., analysing 24 trials and 6007 patients, found that SCT led to significant relapse free survival (RFS) and overall survival (OS) benefits for AML patients with unfavourable genetic mutations and high chance of relapse (8)

Health inequities defined as unfair and avoidable differences in health between social groups, is evidenced by the 8–10-year gap in life expectancy and 20-year gap in healthy life expectancy between the most and least deprived area in the UK (11). Patients living with cancer in deprived areas are 60% more likely to die than those from more affluent areas (12). In the haematological malignancies setting, inequities have been reported to impact on incidence and prevalence of disease, diagnosis, referral, and treatment outcomes. Age-standardised incidence rates of myeloma have been reported to be 2.7-3.0 times higher in people of Black ethnicity compared to white ethnic groups in England (13). Despite this, ethnic minority patients are reported to experience delayed diagnosis and referral (14–16). A survey by the Blood Cancer Alliance showed that 45.5% of ethnic minority patients visited their GP three or more times before being referred compared to 25% of blood cancer patients overall (14,17). An increased incidence of childhood leukaemia in more affluent communities suggests under-diagnosis in the poorest as one of the possible explanations (18,19). Socioeconomic status has been reported to strongly influence outcomes of haematological malignancies. Fegan et al showed significantly better survival in the least deprived group, with a 41% lower risk of mortality compared to the most deprived group of patients with chronic lymphocytic leukeamia (20). Similarly, Bhayat et al, reported poorer survival in AML, with 50% higher mortality rates in patients from the most deprived quintile compared to the least deprived (21).

Despite the survival benefits of SCT, there is growing evidence to suggest that not all patients who could benefit from SCT receive it, with inequities experienced along the SCT treatment pathway, from diagnosis, transplant referral and eligibility assessment to donor identification and transplant (15,22). An inquiry from the All-Party Parliamentary Group (APPG) on SCT access described geographical, cultural, complex health systems, housing, health literacy and education, race, ethnicity, discrimination, and socioeconomic deprivation as barriers faced by patients accessing to SCT treatment and care (15).

Socioeconomic deprivation refers to a lack of income and other resources needed to fully participate in society. Measures of socioeconomic deprivation can be collected at the individual, household (income, education, and occupation) or neighbourhood level using metrics such as crime, housing, local services and living environment (23,24).While socioeconomic deprivation is a key driver of health inequities and a fundamental social determinant of health, modifiable by policy and directly tied to the NHS’s Long-term plan and Core 20plus5 agenda, studies on its role in SCT access and survival have yielded mixed results (25–30). In their 2024 study of AML patients who underwent SCT, Oliveri et al, showed limited impact of socioeconomic and other non-biologic characteristics such as race/ethnicity, and distance travelled on non-relapse mortality (NRM), relapse free survival and OS in multivariate analysis except for marital status with improved NRM (risk of NRM HR = 0.70 (95% CI: 0.50–0.97, p=0.031) and having no insurance with greater risk of death (HR= 1.49 [1.05–2.12, p=0.024) (30). Studies exploring SCT access suggest that poverty, lack of insurance and education have an impact on transplant rates with lower transplant rates described with higher poverty (31–33). The evidence in this area is mostly US based, where the insurance-based health system represents an important barrier to healthcare, contributing to health inequities.

In 2022, the UK Stem cell Strategic forum published a 10-year report for SCT, recommending the need for better understanding of the role of ethnicity and socioeconomic factors on patient access to SCT and outcomes (34). Ethnicity is also a major determinant of health inequity and a well-known barrier to finding an optimal donor source for SCT, especially for patients from an ethnic minority background (35–38). Socioeconomic deprivation and ethnicity are linked and intersect (39). In 2019, people from all ethnic minority groups (except Indian, Chinese, and White Irish) were more likely than White British people to live in the most overall deprived 10% of neighbourhoods in England (40). Ethnicity remains an important area of investigation and while our recent systematic review protocol highlighted the gaps in the UK evidence base of ethnic inequities in SCT, this review will focus on the role of socioeconomic deprivation(41).

### Why it is important to do this review?

There are conflicting results in the literature on the impact of socioeconomic deprivation on SCT access and survival in the haematological malignancy setting. A synthesis of the evidence on the role of socioeconomic deprivation in the SCT treatment pathway in a healthcare system where access is free is important to evaluate the impact of socioeconomic deprivation without the healthcare barriers due to lack of insurance. To our knowledge, no systematic review has been undertaken to synthesise the evidence of the role of socioeconomic deprivation on inequities in the SCT treatment pathway in the UK. This review will focus on studies using UK data only due to differences in societal make up, healthcare infrastructure and drivers of deprivation, which vary from country to country. Identification of the socioeconomic disparities in this UK setting is crucial to help identify inequity in care provision and set policy to address those inequities and improve survival for underserved patients.

### Aims

This systematic review aims to describe the inequities due to socioeconomic deprivation faced by patients at all stages of haematological cancer care for those haematological malignancies treated by SCT, in the UK.

### Objectives of this systematic review

1. Investigate the role of socioeconomic deprivation at different stages of the treatment pathway from the point of diagnosis to treatments/interventions received, referral to transplant, transplant eligibility, transplant utilisation rates, waiting time to transplant and survival rates.
2. Explore any known mechanisms contributing to these inequities.

## Methods and Analysis

Standard systematic review methodology will be used in accordance with the Preferred Reporting Items for Systematic Reviews and Meta-Analyses (PRISMA) guidelines and in line with the Cochrane Handbook (42,43). This protocol is registered with PROSPERO (2024: CRD42024620592) and has been written following PRISMA Protocols and checklist (Supplementary File 1) (44).

### Eligibility Criteria

This review will include all observational studies, including cohort, cross-sectional, case-crossover and case control studies, which examine the impact of socioeconomic deprivation on transplant referral, utilisation, and survival. Qualitative and mixed method research will be excluded due to resource constraints, and we do not anticipate any randomised evidence which will provide insight into this research question. We will include studies which include a UK population, with non-UK populations excluded due to differences in health systems, drivers of socioeconomic deprivation, education, cultures, behaviours, and economic conditions of different nations.

We will include individual-level and household socio-economic determinants such as income, education, employment status, poverty/low wage, social isolation, car ownership and housing status and aggregate area-level material and relative measures including the Index of Multiple Deprivation (IMD), Townsend Deprivation Index, and Carstairs Deprivation Index (45).

All inclusion and exclusion criteria are detailed in Table 1.

**Table 1.**
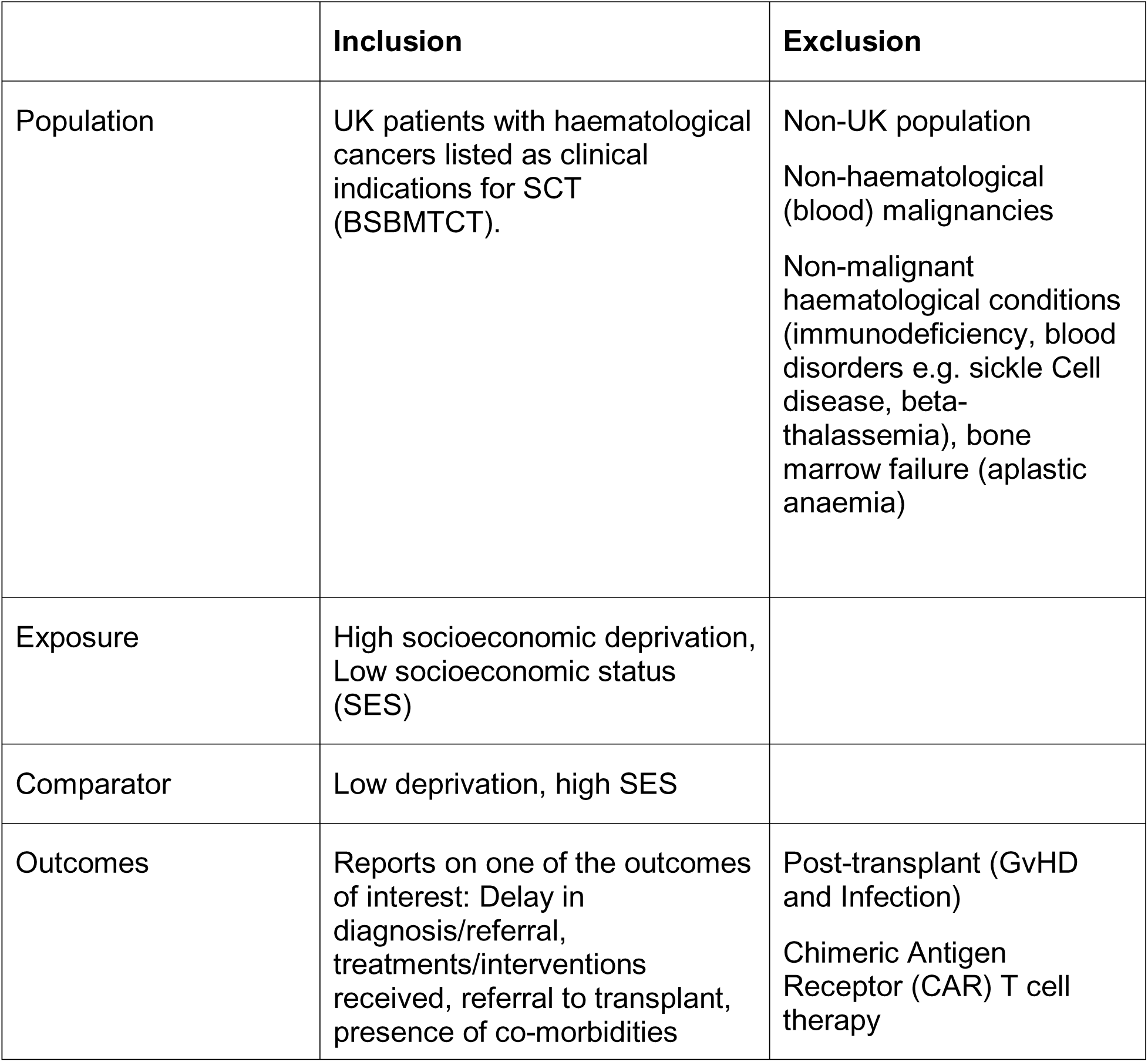
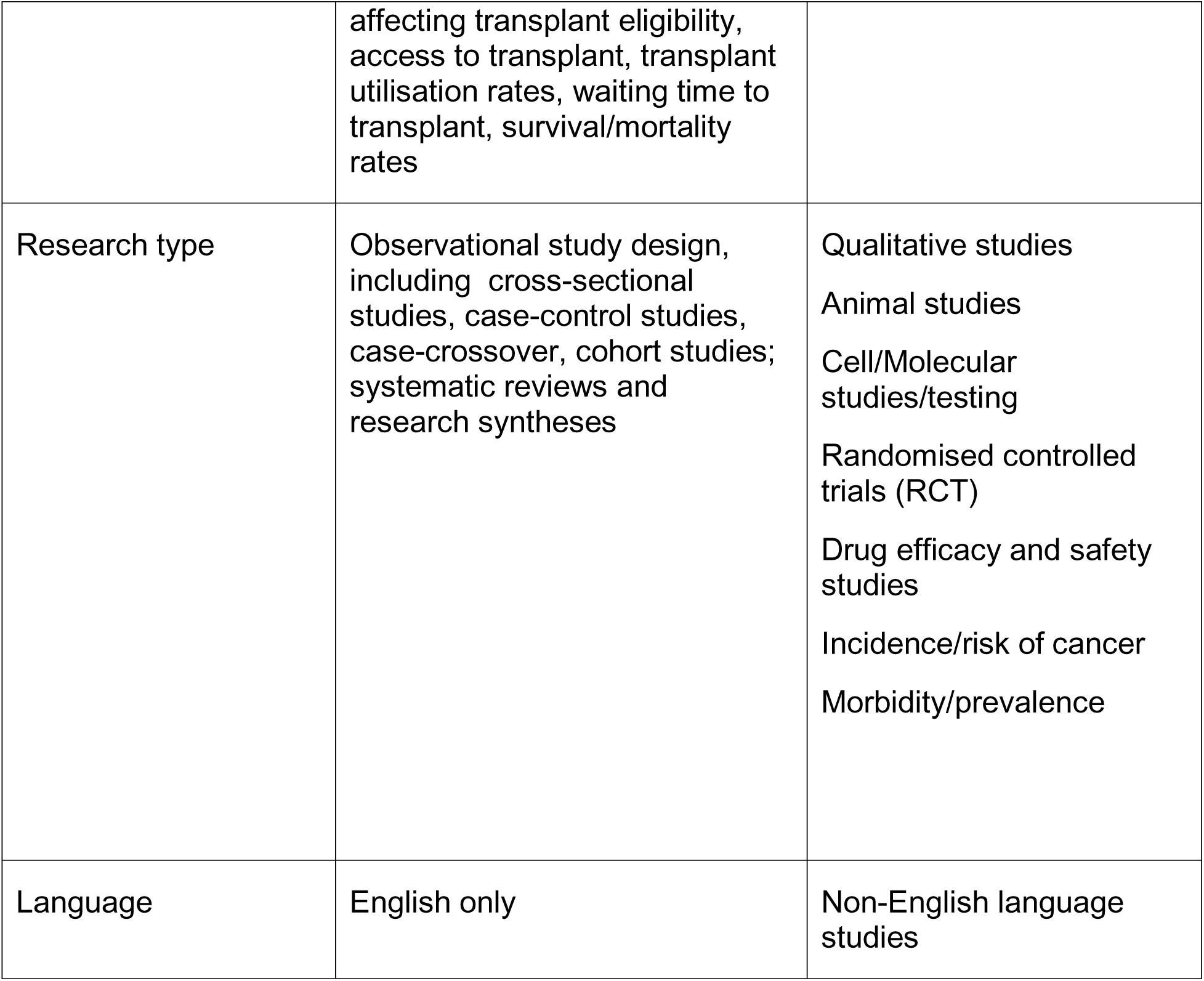
Inclusion and exclusion criteria.

|  | **Inclusion** | **Exclusion** |
| --- | --- | --- |
| Population | UK patients with haematological cancers listed as clinical indications for SCT (BSBMTCT). | Non-UK population<br>Non-haematological (blood) malignancies<br>Non-malignant haematological conditions (immunodeficiency, blood disorders e.g. sickle Cell disease, beta-thalassemia), bone marrow failure (aplastic anaemia) |
| Exposure | High socioeconomic deprivation, Low socioeconomic status (SES) |  |
| Comparator | Low deprivation, high SES |  |
| Outcomes | Reports on one of the outcomes of interest: Delay in diagnosis/referral, treatments/interventions received, referral to transplant, presence of co-morbidities | Post-transplant (GvHD and Infection)<br>Chimeric Antigen Receptor (CAR) T cell therapy |
|  | affecting transplant eligibility, access to transplant, transplant utilisation rates, waiting time to transplant, survival/mortality rates |  |
| Research type | Observational study design, including cross-sectional studies, case-control studies, case-crossover, cohort studies; systematic reviews and research syntheses | Qualitative studies<br>Animal studies<br>Cell/Molecular studies/testing<br>Randomised controlled trials (RCT)<br>Drug efficacy and safety studies<br>Incidence/risk of cancer<br>Morbidity/prevalence |
| Language | English only | Non-English language studies |

UK patients with haematological cancers which are clinical indications for SCT as listed by the BSBMTCT will be included (46) This classification of haematological cancers was used due to the focus of this review on a UK population. These haematological malignancies are:

- Leukaemia

- Chronic Myeloid Leukaemia (CML)
- Acute Myeloid Leukaemia (AML)
- Acute Lymphoblastic Leukaemia (ALL)
- Lymphomas

- Hodgkin’s Lymphoma

- Classical Hodgkin’s Lymphoma
- Nodular Lymphocyte-Predominant B-cell Lymphoma
- Non-Hodgkin’s Lymphomas

- Peripheral T cell lymphoma (PTCL)
- Diffuse large B-cell lymphoma (DLBCL)
- Mantle cell Lymphoma
- Follicular Lymphoma
- Burkitt lymphoma
- Marginal zone lymphoma (MCL)
- Primary CNS lymphoma
- Extranodal marginal zone lymphoma of mucosa-associated lymphoid tissue (MALT) lymphoma
- Acute Lymphoblastic Lymphoma (ALL)
- Lymphoplasmacytic Lymphoma
- Waldenström’s macroglobulinaemia
- Chronic Lymphocytic Leukaemia (CLL)/Small Lymphocytic Lymphoma (Richter’s syndrome)
- Plasma cell dyscrasias

- Myeloma (Multiple Myeloma (MM)
- AL amyloid (Amyloid light chain (AL) amyloidosis)
- POEMS (Polyneuropathy, organomegaly, endocrinopathy, monoclonal gammopathy, and skin changes) syndrome
- Myelofibrosis (primary/secondary)
- Myelodysplastic syndrome (myelodysplasia, myelodysplastic neoplasm) (MDS)

### Information Sources

The following databases and search tools will be searched for relevant evidence: MEDLINE, Embase, Proquest Databases, CENTRAL & CDSR (*The Cochrane Library),* Stem Cell Evidence, Web of Science, ClinicalTrials.gov, WHO International Clinical Trials Registry Platform (ICTRP), Google Scholar will be also searched to provide grey literature coverage (first 10 pages of results).

Additionally, we used Google Scholar’s advanced search feature using the search string detailed below in the ‘Search Strategy’ section to identify grey literature listed in English.

We will also manually check references lists and citations of eligible studies to capture additional data to ensure full coverage of eligible literature.

### Search Strategy

A search strategy (Table 2; Supplementary File 2) has been developed by co-author CD broadly including the terms “Socioeconomic Factors”, “health Inequities”, “health Services Accessibility”, “Social Determinants of Health”, “Social Conditions”, “Social Socioeconomic deprivation”, “Social Environment”, “Sociodemographic Factors”, “Social Isolation”, “Social Marginalization”, “Social Vulnerability”, “Stem Cell Transplantation”, Bone Marrow Transplantation”, “Cell Transplantation”, “Hematologic Neoplasms”, “Lymphoma, Leukemia”, “Neoplasms”, “Plasma Cell”, “Bone Marrow Diseases”, “Great Britain” and “United Kingdom”.

Studies in languages other than English will be excluded due to resource constraints.

**Table 2.**
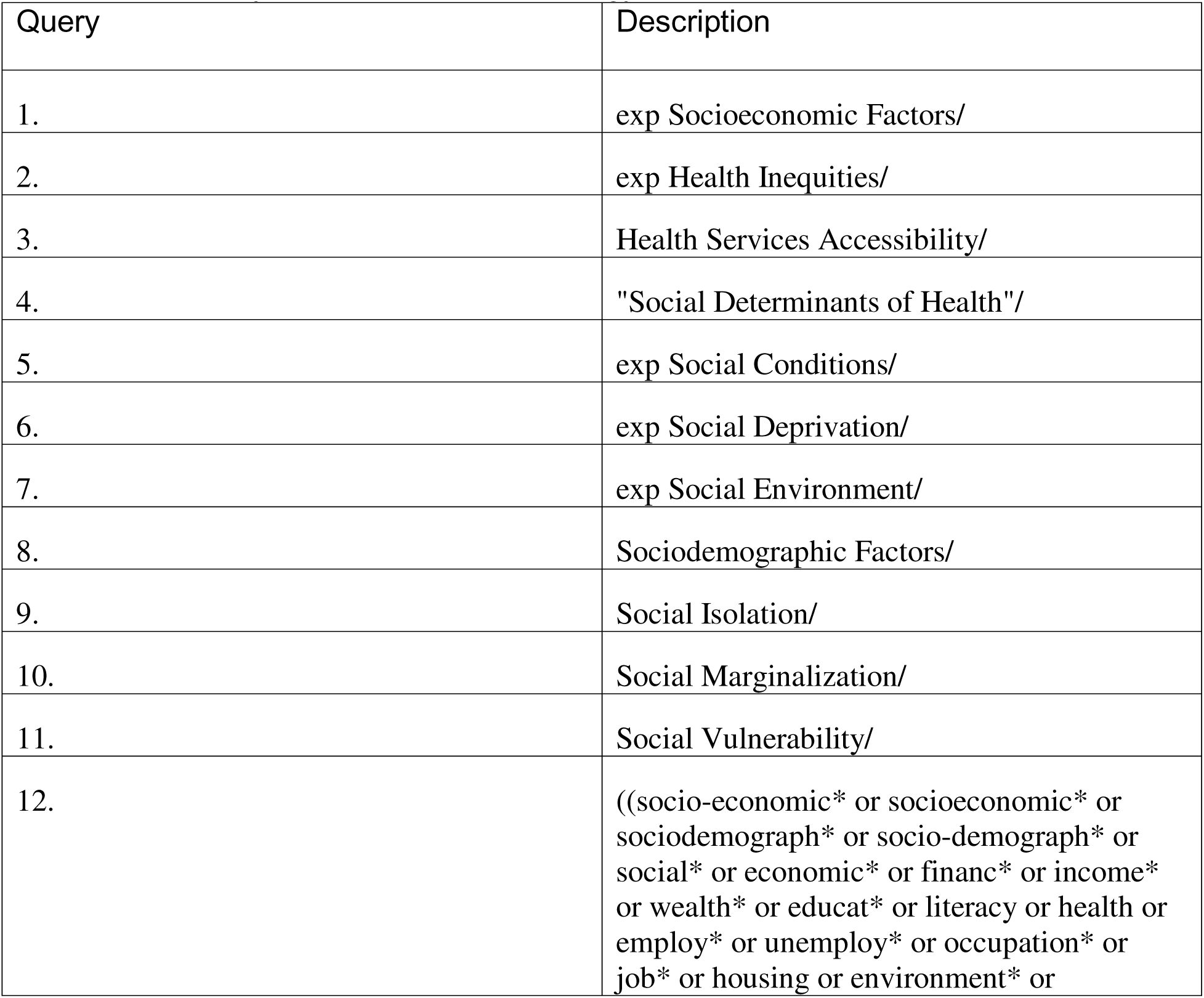
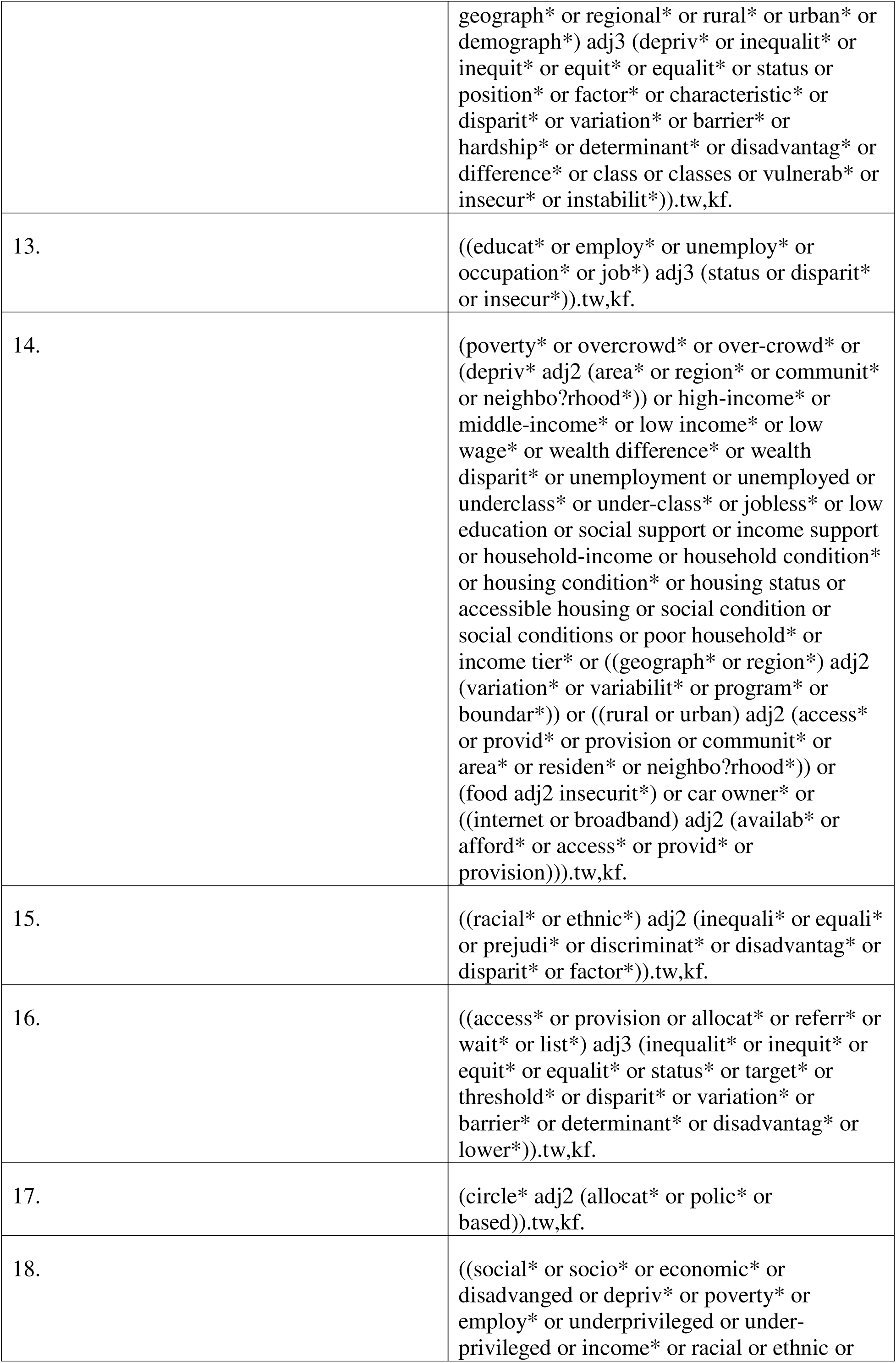
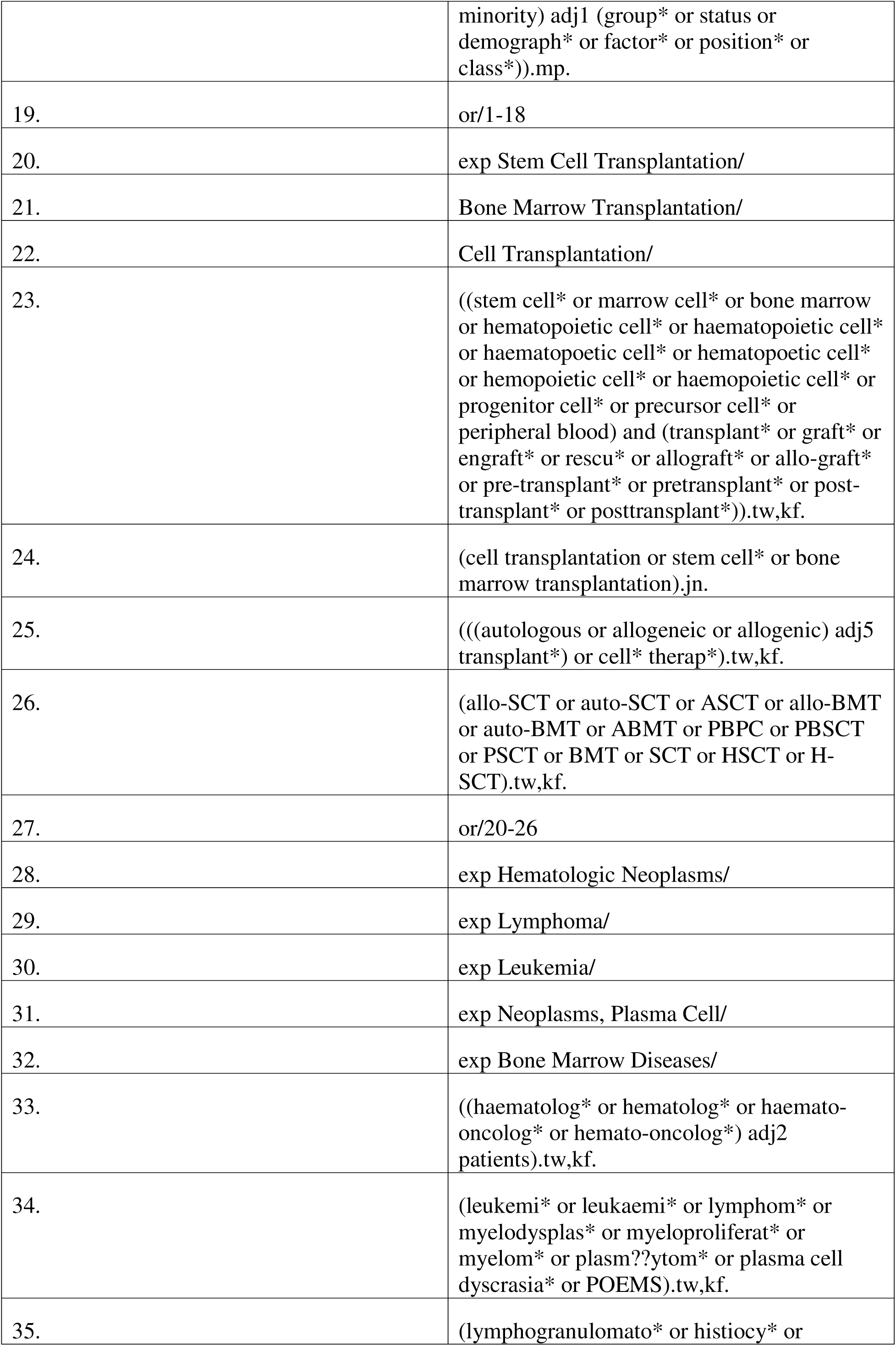
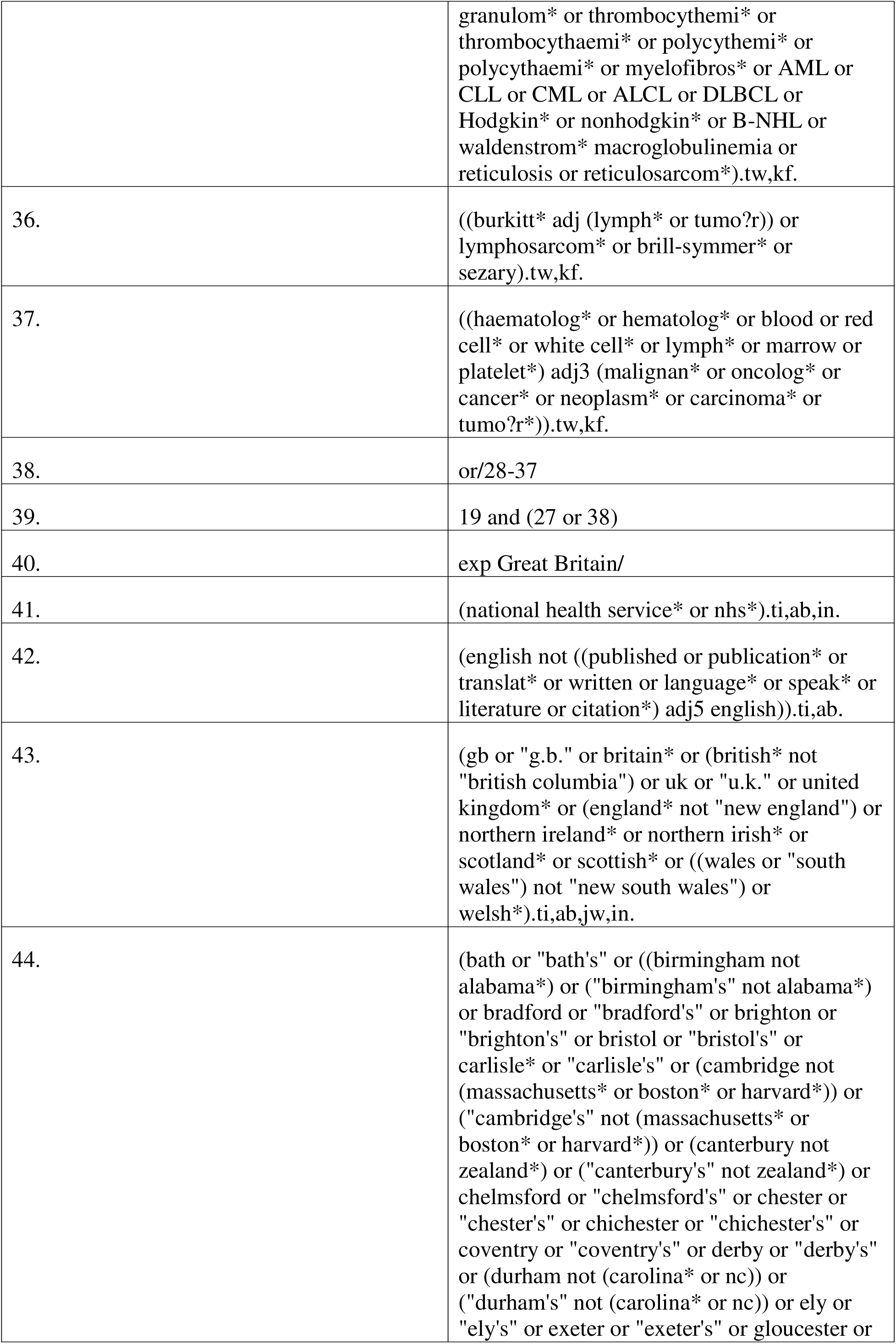
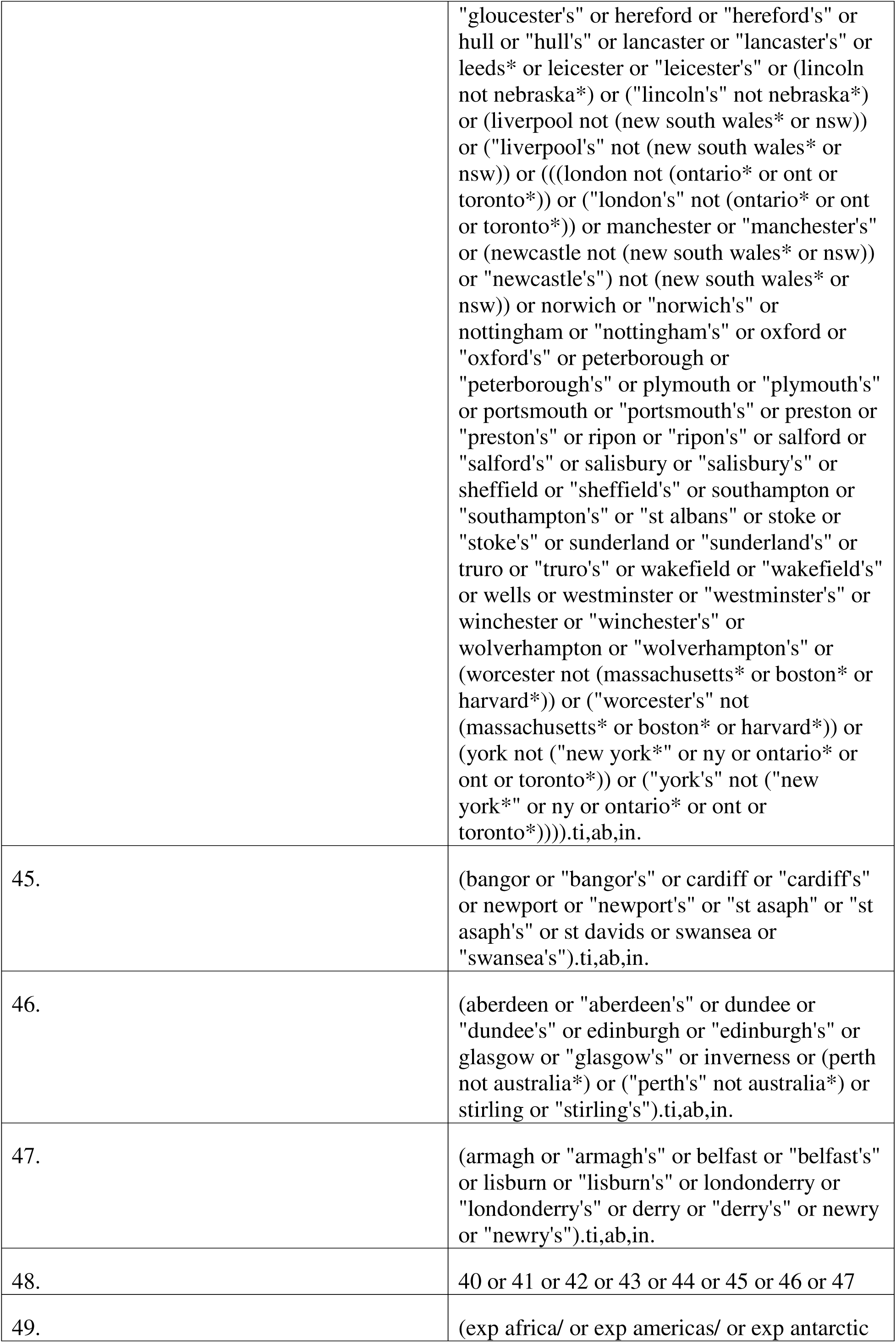
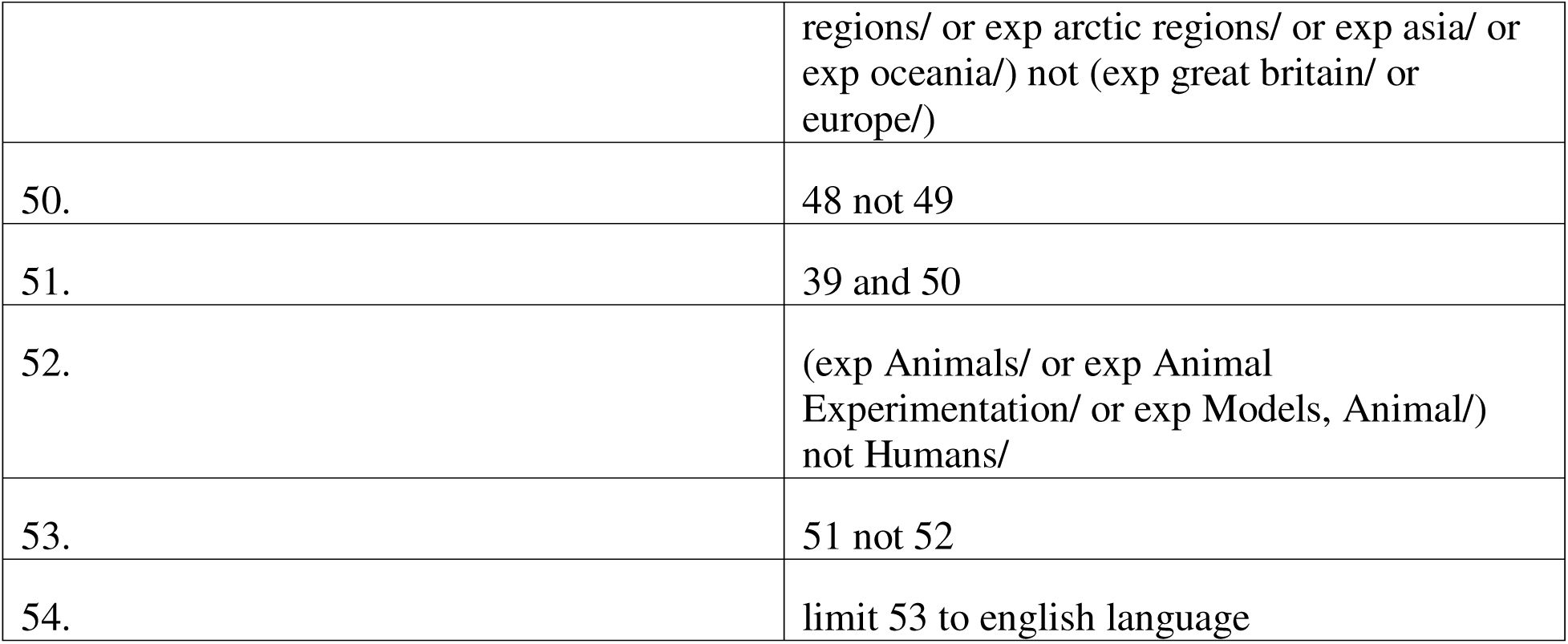
Summary of Medline search strategy.

Google scholar Search string: ((deprivation OR socioeconomic) OR (determinants AROUND (1) health) OR (inequities OR inequalities OR access)) AND ((((haematological OR blood) AROUND (1) (cancer OR malignancies)) OR lymphoma OR leukaemia OR myeloma OR myelodysplastic) OR ((“bone marrow transplant” OR “stem cell transplant” OR “haematopoietic cell transplant”) OR (BMT OR SCT OR HSCT))) AND (“united kingdom” OR england OR uk OR britain)

### Study Records and Data Management

Covidence will be used to conduct and organise reviewing of the searched evidence, including removal of duplicates (47). Mendeley Reference Manager will be used to organise the literature extracted after full text review (48). This will also be used to automate the downloading of open-access papers. The PRISMA flow diagram will be used to summarise the screening and selection of the studies (49).

### Study Selection Process

Two independent reviewers will conduct title and abstract screening according to the pre-defined inclusion/exclusion criteria (Table 1). Any disagreement unresolved through discussion will be evaluated by a third party (JSC or NJA). Full texts of the shortlisted articles will be obtained and exclusion criteria applied by two reviewers (ZD, SC) based on the above inclusion and exclusion criteria, with an additional reviewer providing support to resolve any disagreements ((JSC or NJA). The final list of eligible studies will be obtained for inclusion in the review after the full text review.

### Data collection

The identified studies meeting the eligibility criteria will be inputted into the data extraction form, which uses a modified Cochrane Public Health Group Data Extraction and Assessment Template. The data extraction form will be piloted by two reviewers to provide an opportunity to refine the data extraction process and ensure the relevant data items are being collected (Supplementary File 3). Data extraction will be done in independently, in duplicate, by ZD and SC.

Any discrepancies on selection and extraction will be resolved through discussion between the reviewers and the project leads (JC and NJA).

### Data items

For each included article, the following data items will be extracted based on the study Population Exposure Comparator Outcome: 1) Study information (Author, year of publication, Conflict of interest, ethical approval), 2) Study characteristics (Study design and type, duration, data source, aim, recruitment, statistical analysis 3) Population Characteristics (description of main characteristics, treatment, co-morbidities) 4) Exposure and outcomes (measurement of socioeconomic deprivation at individual, neighbourhood or aggregate level e.g. IMD, Carstairs, Townsend Index) 5) Effect size and uncertainty 6) Main Findings, 7) Key conclusions (Supplementary File 3).

### Risk of Bias and Quality assessment in Individual Studies

The risk of bias for the included studies will be assessed by 2 reviewers (ZD and SC) using the Newcastle-Ottawa Scale (NOS) which assesses the quality of non-randomised studies based on their selection, comparability, and outcome/exposure characteristics. High quality choices within each domain are awarded stars which are converted to ratings: good, fair, and poor, which will be recorded on the data extraction form. The rating will not lead to any study being excluded, but caution will be used during data analysis for studies rated poor and fair quality.

Any discrepancies on selection and extraction will be resolved through discussion between the reviewers and the project leads (JC and NJA).

### Outcomes and prioritization

Main outcomes:

1. SCT transplant referral (referral to a specialist tertiary centre/transplant centre), including SCT referral rates %, number of SCT referrals, likelihood of SCT referral OR, 95%CI, stratified according to socioeconomic deprivation;
2. SCT utilisation/rate, including transplant rate %, number of SCT, likelihood of SCT, stratified according to socioeconomic deprivation.;
3. Access to SCT (referred to transplant), including numbers referred, referral rate, referral patterns, likelihood of referral OR, 95% CI), assessed for transplant eligibility, receipt of transplant (number of transplants received);
4. Survival, including overall survival/mortality, transplant survival rates, % survival.

Additional outcomes of interest include:

1. Delayed diagnosis measured by multiple GP visits/multiple symptoms (number or %), place of presentation (presents with symptoms at GP, through other investigations, symptoms require A&E admission, place of diagnosis, number of diagnosis in the following setting A&E, GP, secondary care, advanced disease/ genetic risk/grade of disease at presentation);
2. Treatments/interventions received (number or % patients receiving intensive chemotherapy/conditioning, systemic anti-cancer therapy);
3. Time between different stages of the treatment pathway;
4. Presence of co-morbidities (Charlson Comorbidity index (CI), Hematopoietic cell transplantation-specific comorbidity index (HCTCI) scoring systems used by Transplant Centres).

### Data Synthesis

Narrative synthesis will be conducted on the findings from the review, where outcomes include delay in diagnosis/referral treatments/interventions received, referral to transplant, presence of co-morbidities affecting transplant eligibility, access to transplant, transplant utilisation rates, waiting time to transplant survival rates will be presented under broad headings with detailed summary of findings.

If data is available and homogeneous and there are at least three studies with comparable measurement of socio-economic deprivation (individual, household, aggregate) reporting the outcomes of interest, meta-analysis will be conducted. Only studies using the same study designs will be considered for pooling. Meta-analysis of Observational Studies in Epidemiology (MOOSE) recommendations will be used to guide the meta-analysis of observational studies (50).

We will produce pooled odds ratios using a random or fixed-effect model to assess likelihood of transplant referral and/or utilisation according to measures of socioeconomic deprivation (least deprived/most deprived).

Within each outcome, meta-analysis will follow for each individual-level, household, or aggregate socioeconomic determinant exposure if there are three or more studies reporting sufficiently similar combinations of exposures and outcomes. Summaries of exposure effects for each study will be provided by calculating a pooled odds ratio (OR), hazard ratio (HR), risk ratio (RR) and risk difference (RD). Tests will be applied for heterogeneity between included studies using the I² test statistic.

In absence of heterogeneity in at least 10 studies, publication bias will be assessed using the Egger’s regression test.

### Confidence in cumulative evidence

The certainty of evidence for each outcome will be assessed using the Grades Recommendations, Assessment, Development and Evaluation (GRADE) approach. As most included studies are expected to be observational (registry-based or cohort), evidence will begin at a low certainty rating. Certainty may be downgraded for risk of bias (e.g., incomplete adjustment for confounders), inconsistency (variation in effect estimates), indirectness (differences in definitions of access or outcomes), imprecision (wide confidence intervals or small samples), or publication bias. Where appropriate, evidence may be upgraded if there is a large, consistent effect, a dose–response gradient, or if residual confounding would likely reduce the observed association.

Certainty of evidence will be graded as high, moderate, low, or very low for each primary and additional outcome (Delay in diagnosis/referral, treatments/interventions received, referral to transplant, presence of co-morbidities affecting transplant eligibility, access to transplant, transplant utilisation rates, waiting time to transplant and survival rates) (51)

### Measurements of treatment effect

For binary data, pooled OR with 95% CIs or other effect estimates will be presented. For continuous data, we will use the mean difference with 95% CIs.

### Subgroup analysis

If data is available, analysis of inequities at different points in the SCT treatment pathway for haematological malignancies will be conducted.

## Patient and public involvement

Patients and the public participated in the development of the review. This was undertaken with the support of a patient and public involvement and engagement (PPIE) Steering Committee made up of six representatives from transplant charities (Team Margo, African Caribbean Leukaemia Trust (ACLT), research groups and stem cell donor registry in the UK (NHSBT, Anthony Nolan). There was a mix of ethnic groups and experience represented among the members including a patient and two caregivers. Following the presentation of the rationale and preliminary ideas for this review to the group, the scope and objectives of the review were discussed. While the PPIE steering committee thought the objectives and scope were appropriate, they emphasised the need to explore the link between ethnicity and socioeconomic deprivation. Based on their feedback, we will ensure that this review also reports outcomes where the impact of socioeconomic deprivation on the blood cancer patient pathway is described within different ethnic minority groups if this data is available. Patients and members of the public will not be involved in the study selection or extraction phases but will be involved in the evidence synthesis stage of the review and this will be documented according to Patient Student Partner (PSP) Activity log. The final review and lay summary will be supported by the PPIE Steering Committee and disseminated via publications in scientific journals, conference presentations and to the wider community at patient facing meetings such NBTA and the BTRU.

## Data Availability

All data produced in the present work are contained in the manuscript

## Author Statement

The research questions were conceived by ZD, JSC, NJA, JG. The first draft of the protocol was done by ZD, JSC, NJA, JG. Further reviews and revisions to the protocol were made by all co-authors (ZD, JSC, NJA, JG SC). All authors approved the final copy of the manuscript.

The funder was not involved in the development of this protocol.

ZD is the guarantor

## Patient and public involvement

Patients and/or the public participated in the design, or conduct, or reporting, or dissemination plans of this research. Refer to the Methods section for further details.

Kirsty Brown for critical review of the draft protocol.

## Ethics approval

This study does not require ethical approval as it is a secondary analysis of published data.

## References

1. Haematological Malignancy Research Network (HMRN). HMRN. 2017 [cited 2025 Sep 4]. HMRN - Haematological malignancies. Available from: https://hmrn.org/about/classification

2. Jurlander J. Hematological Malignancies, Leukemias and Lymphomas. Encyclopedia of Cancer [Internet]. 2008 Oct 16 [cited 2025 Sep 4];1328–32. Available from: https://link.springer.com/rwe/10.1007/978-3-540-47648-1_2615

3. ONS. ONS. [cited 2024 Apr 17]. Cancer registration statistics, England - Office for National Statistics. Available from: https://www.ons.gov.uk/peoplepopulationandcommunity/healthandsocialcare/conditionsanddiseases/bulletins/cancerregistrationstatisticsengland/2017

4. Blood Cancer UK. Blood cancer facts | Blood Cancer UK [Internet]. [cited 2024 Jun 6]. Available from: https://bloodcancer.org.uk/news/blood-cancer-facts/

5. National Disease Registration Service. Get Data Out [Internet]. NHS England; 2023 [cited 2025 Sep 18]. Available from: https://nhsd-ndrs.shinyapps.io/get_data_out/

6. Loke J, Buka R, Craddock C. Allogeneic stem cell transplantation for acute myeloid leukemia: who, when, and how? Front Immunol [Internet]. 2021 [cited 2025 Sep 18];12:659595. Available from: https://www.frontiersin.org/articles/10.3389/fimmu.2021.659595/full

7. Cavo M, Gay F, Beksac M, Dimopoulos MA, Pantani L, Petrucci MT, et al. Upfront Autologous Hematopoietic Stem-Cell Transplantation Improves Overall Survival in Comparison with Bortezomib-Based Intensification Therapy in Newly Diagnosed Multiple Myeloma: Long-Term Follow-up Analysis of the Randomized Phase 3 EMN02/HO95 Study. Blood [Internet]. 2020 Nov 5 [cited 2025 Jun 17];136(Supplement 1):37–8. Available from: https://www.sciencedirect.com/science/article/pii/S0006497118693632

8. Koreth J, Schlenk R, Kopecky KJ, Honda S, Sierra J, Djulbegovic BJ, et al. Allogeneic stem cell transplantation for acute myeloid leukemia in first complete remission: a systematic review and meta-analysis of prospective clinical trials. JAMA. 2009;301(22):2349–61. Available from: https://jamanetwork.com/journals/jama/fullarticle/184068

9. BSBMTCT, Activity report 2023. BSBMTCT. [cited 2025 Jan 23]. 2023 - British Society of Blood and Marrow Transplantation. Available from: https://bsbmtct.org/activity/2023/

10. Cornelissen JJ, Breems D, van Putten WLJ, Gratwohl A, Passweg JR, Pabst T, et al. Comparative analysis of the value of allogeneic hematopoietic stem-cell transplantation in acute myeloid leukemia with monosomal karyotype versus other cytogenetic risk categories. J Clin Oncol [Internet]. 2012 [cited 2025 Sep 18];30(17):2140–6. Available from: https://ascopubs.org/doi/full/10.1200/JCO.2011.39.6499

11. Office for National Statistics. Healthy life expectancy by national area deprivation, England and Wales - Office for National Statistics [Internet]. 2025 [cited 2025 Sep 16]. Available from: https://www.ons.gov.uk/peoplepopulationandcommunity/healthandsocialcare/healthinequalities/bulletins/healthylifeexpectancybynationalareadeprivationenglandandwales/between2013to2015and2020to2022

12. O’Dowd A. Cancer: people in UK’s poorest areas have longer waits for care and higher death rates. BMJ [Internet]. 2025 Feb 21 [cited 2025 Sep 18];388:r367. Available from: https://www.bmj.com/content/388/bmj.r367

13. Delon C, Brown KF, Payne NWS, Kotrotsios Y, Vernon S, Shelton J. Differences in cancer incidence by broad ethnic group in England, 2013–2017. Br J Cancer [Internet]. 2022 Jun 1 [cited 2025 Sep 18];126(12):1765–73. Available from: https://www.nature.com/articles/s41416-022-01718-5

14. Blood Cancer UK. Taking blood cancer out of the shadows: a plan to increase survival in the UK. Blood Cancer UK [Internet]. 2024 Sep 4 [cited 2025 Sep 18]. Available from: https://bloodcancer.org.uk/news/uk-blood-cancer-action-plan-launched

15. Anthony Nolan, The Secretariat to the APPG on Stem Cell Transplantation. No patient left behind: the barriers stem cell transplant patients face when accessing treatment and care. London: Anthony Nolan; 2021 [cited 2025 Sep 18]. Available from: https://www.anthonynolan.org/sites/default/files/2021-05/no_patient_left_behind_final.pdf

16. Herbert A, Abel GA, Winters S, McPhail S, Elliss-Brookes L, Lyratzopoulos G. Cancer diagnoses after emergency GP referral or A&E attendance in England: determinants and time trends in routes to diagnosis data, 2006–2015. Br J Gen Pract [Internet]. 2019 [cited 2025 Sep 18];69(687):e724–30. Available from: https://bjgp.org/content/69/687/e724

17. Hicks S, McGarry N. Blood cancer patient experience for Black, Asian and ethnic minority patients. ClearView Research [Internet]. 2022 [cited 2025 Sep 18]. Available from: https://static1.squarespace.com/static/5b98cdc612b13fdd2982129d/t/6387561da2a03105934f1bad/1669813794902/Blood+Cancer+Patient+Experience+for+Black%2C+Asian+and+Ethnic+Minority+Patients.pdf

18. Kroll ME, Stiller CA, Murphy MFG. Reply: Childhood leukaemia and socioeconomic status. Br J Cancer [Internet]. 2012 Jun 12 [cited 2025 Sep 18];107:216. Available from: https://www.nature.com/articles/bjc2012171.pdf

19. Kroll ME, Stiller CA, Murphy MFG, Carpenter LM. Childhood leukaemia and socioeconomic status in England and Wales 1976–2005: evidence of higher incidence in relatively affluent communities persists over time. Br J Cancer [Internet]. 2011 Nov 22 [cited 2025 Sep 18];105(11):1783–7. Available from: https://www.nature.com/articles/bjc2011415.pdf

20. Fegan G, Cashell R, DTADNECP. Social deprivation independently impacts clinical outcomes in patients with chronic lymphocytic leukemia. Haematologica [Internet]. 2024 [cited 2025 Sep 18];109(6): 1566–9. Available from: https://haematologica.org/article/view/haematol.2023.283527

21. Bhayat F, Das-Gupta E, Smith C, McKeever T, Hubbard R. The incidence of and mortality from leukaemias in the UK: a general population-based study. BMC Cancer [Internet]. 2009 Jul 26 [cited 2025 Sep 18];9:252. Available from: https://bmccancer.biomedcentral.com/articles/10.1186/1471-2407-9-252

22. Eapen M. Since everyone has a donor, why are some eligible patients still not transplanted? Best Pract Res Clin Haematol [Internet]. 2021 [cited 2024 Apr 18];34:1521–6926. Available from: 10.1016/j.beha.2021.101321

23. Mack J. Deprivation and poverty | Poverty and Social Exclusion. UKRI Economic and Social Research Council [Internet]. 2016 [cited 2025 Sep 18]. Available from: https://www.poverty.ac.uk/definitions-poverty/deprivation-and-poverty

24. Ministry of Housing, Communities & Local Government. English indices of deprivation 2019: research report. GOV.UK [Internet]. 2019 [cited 2025 Sep 18]. Available from: https://www.gov.uk/government/publications/english-indices-of-deprivation-2019-research-report

25. NHS England. The NHS Long Term Plan. NHS [Internet]. 2019 [cited 2025 Sep 18]. Available from: https://www.longtermplan.nhs.uk

26. NHS England » Core20PLUS5 – An approach to reducing health inequalities for children and young people [Internet]. [cited 2024 Jan 22]. Available from: https://www.england.nhs.uk/about/equality/equality-hub/national-healthcare-inequalities-improvement-programme/core20plus5/core20plus5-cyp/

27. Baker KS, Davies SM, Majhail NS, Hassebroek A, Klein JP, Ballen KK, et al. Race and socioeconomic status influence outcomes of unrelated donor hematopoietic cell transplantation. Biol Blood Marrow Transplant [Internet]. 2009 Dec [cited 2025 Sep 18];15(12):1543–54. Available from: https://europepmc.org/articles/PMC2775819

28. Bona K, Brazauskas R, He N, Lehmann L, Abdel-Azim H, Ahmed IA, et al. Neighborhood poverty and pediatric allogeneic hematopoietic cell transplantation outcomes: a CIBMTR analysis. Blood [Internet]. 2021 Jan 28 [cited 2025 Sep 18];137(4):556–68. Available from: https://ashpublications.org/blood/article/137/4/556/469714

29. Garcia L, Feinglass J, Marfatia · Hardik, Kehinde Adekola ·, Moreira J, Marfatia H, et al. Evaluating Socioeconomic, Racial, and Ethnic Disparities in Survival Among Patients Undergoing Allogeneic Hematopoietic Stem Cell Transplants. J Racial Ethn Health Disparities [Internet]. 2023 [cited 2024 Apr 22]; Available from: 10.1007/s40615-023-01611-8

30. Olivieri DJ, Othus M, Orvain C, Rodríguez-Arbolí E, Milano F, Sandmaier BM, et al. Impact of socioeconomic disparities on outcomes in adults undergoing allogeneic hematopoietic cell transplantation for acute myeloid leukemia. Leukemia [Internet]. 2024 [cited 2025 Sep 18];38:865–76. Available from: 10.1038/s41375-024-02172-3

31. Paulson K, Brazauskas R, Khera N, He N, Majhail N, Akpek G, et al. Quality of care: inferior access to allogeneic transplant in disadvantaged populations—a Center for International Blood and Marrow Transplant Research analysis. Biol Blood Marrow Transplant [Internet]. 2019 [cited 2025 Sep 18];25(10):2086–90. Available from: 10.1016/j.bbmt.2019.06.012

32. Bhatt VR, Chen B, Lee SJ. Use of hematopoietic cell transplantation in younger patients with acute myeloid leukemia: a National Cancer Database study. Bone Marrow Transplant [Internet]. 2018 [cited 2025 Sep 18];53:873–9. Available from: 10.1038/s41409-018-0105-9

33. Jabo B, Morgan JW, Martinez ME, Ghamsary M, Wieduwilt MJ. Sociodemographic disparities in chemotherapy and hematopoietic cell transplantation utilization among adult acute lymphoblastic and acute myeloid leukemia patients. PLoS One [Internet]. 2017 [cited 2025 Sep 18];12(3):e0174760. Available from: 10.1371/journal.pone.0174760

34. Cradock C, Miflin G, UK Stem Cell Strategic Forum. 10 year strategy for stem cell transplantation and cellular therapies. NHS Blood and Transplant [Internet]. 2022 [cited 2025 Sep 18]. Available from: https://nhsbtdbe.blob.core.windows.net/umbraco-assets-corp/28283/a-10-year-vision-for-hsct-and-cellular-therapies-august-2022-8.pdf

35. Lown RN, Marsh SGE, Blake H, Querol S, Evseeva I, Mackinnon S, et al. Equality of access to transplant for ethnic minority patients through use of cord blood and haploidentical transplants. Blood [Internet]. 2013 [cited 2025 Sep 18];122(21):2138. Available from: https://ashpublications.org/blood/article/122/21/2138/12152

36. Lown RN, Shaw BE. Beating the odds: factors implicated in the speed and availability of unrelated haematopoietic cell donor provision. Bone Marrow Transplant [Internet]. 2013 [cited 2025 Sep 18];48:210–9. Available from: https://www.nature.com/articles/bmt201254.pdf

37. Lown RN, Marsh SG, Blake H, Querol S, Evseeva I, Mackinnon S, et al. Equality of access to transplant for ethnic minority patients through use of cord blood and haploidentical transplants. Blood [Internet]. 2013 Nov 15 [cited 2025 Sep 18];122(21):2138. Available from: https://ashpublications.org/blood/article/122/21/2138/12152

38. Auletta JJ. Health inequity has no boundaries. Lancet Haematol [Internet]. 2024 Dec 12 [cited 2025 Sep 18]. Available from: https://www.thelancet.com/journals/lanhae/article/PIIS2352-3026(24)00316-8/abstract

39. Coates MJP, Bhardwaj SA, Igwe R, Wan YI. The relationship between ethnicity and socioeconomic deprivation as determinants of health: a systematic review. Epidemiol Public Health [Internet]. 2024 Feb 16 [cited 2025 Sep 18];2(1):1030. Available from: https://jpublichealth.org/articles/1030.html

40. GOV.UK. People living in deprived neighbourhoods - GOV.UK Ethnicity facts and figures [Internet]. 2020 [cited 2025 Jan 21]. Available from: https://www.ethnicity-facts-figures.service.gov.uk/uk-population-by-ethnicity/demographics/people-living-in-deprived-neighbourhoods/latest/

41. Cusworth S, Deplano Z, Denniston A, Burns D, Nirantharakumar K, Adderley NJ, et al. Protocol: what are the ethnic inequities in care outcomes related to haematological malignancies, treated with transplant/cellular therapies, in the UK? A systematic review. BMJ Open [Internet]. 2025 May 17 [cited 2025 Sep 19];15(5):e099354. Available from: https://bmjopen.bmj.com/content/15/5/e099354

42. Page MJ, McKenzie JE, Bossuyt PM, Boutron I, Hoffmann TC, Mulrow CD, et al. The PRISMA 2020 statement: an updated guideline for reporting systematic reviews. BMJ [Internet]. 2021 Mar 29 [cited 2024 Jun 6];372:n71. Available from: 10.1136/bmj.n71

43. Cochrane. Cochrane Handbook for Systematic Reviews of Interventions | Cochrane Training [Internet]. [cited 2024 Apr 9]. Available from: https://training.cochrane.org/handbook/current

44. Moher D, Shamseer L, Clarke M, Ghersi D, Liberati A, Petticrew M, et al. Preferred reporting items for systematic review and meta-analysis protocols (PRISMA-P) 2015 statement. Syst Rev [Internet]. 2015 Jan 1 [cited 2024 Jun 6];4(1):1. Available from: 10.1186/2046-4053-4-1

45. Clinical Practice Research Datalink (CPRD). Small area level data based on patient postcode [Internet]. London: Medicines and Healthcare products Regulatory Agency; 2024 [cited 2025 Sep 18]. Available from: https://www.cprd.com/data/linked-data/small-area-level-data

46. British Society of Blood and Marrow Transplantation and Cellular Therapy (BSBMTCT). HSCT Adult Indications [Internet]. London: BSBMTCT; 2023 [cited 2025 Sep 18]. Available from: https://bsbmtct.org/indications-sub-committee/

47. Covidence. Covidence - Better systematic review management [Internet]. [cited 2024 Jun 6]. Available from: https://www.covidence.org/

48. Elsevier. Mendeley Reference Manager [Internet]. [cited 2024 Jun 6]. Available from: https://www.mendeley.com

49. Page MJ, McKenzie JE, Bossuyt PM, Boutron I, Hoffmann TC, Mulrow CD, et al. PRISMA 2020 flow diagram for new systematic reviews [Internet]. PRISMA Statement; 2021 [cited 2025 May 25]. Available from: https://www.prisma-statement.org/prisma-2020-flow-diagram

50. Stroup DF, Berlin JA, Morton SC, Olkin I, Williamson GD, Rennie D, et al. Meta-analysis of Observational Studies in Epidemiology: A Proposal for Reporting. JAMA [Internet]. 2000 Apr 19 [cited 2024 Jul 8];283(15):2008–12. Available from: https://jamanetwork.com/journals/jama/fullarticle/192614

51. Guyatt GH, Oxman AD, Vist GE, Kunz R, Falck-Ytter Y, Alonso-Coello P, et al. GRADE: an emerging consensus on rating quality of evidence and strength of recommendations. BMJ. 2008 Apr 26;336(7650):924–6. Available from: https://www.bmj.com/content/336/7650/924

