## Supplementary File 2 for "The role of socioeconomic deprivation on inequities in care outcomes of haematological malignancies, treated with stem cell transplant in the UK: A systematic Review Protocol"

**THE ROLE OF DEPRIVATION ON INEQUITIES IN CARE OUTCOMES OF HAEMATOLOGICAL MALIGNANCIES TREATED WITH STEM CELL TRANSPLANT IN THE UK: A SYSTEMATIC REVIEW - SEARCH NARRATIVE, AUGUST 2024**

The following databases were searched for all types of studies on 27.8.24:

- MEDLINE (Ovid, 1946 onwards)
- Embase (Ovid, 1974 onwards)
- Proquest Databases:
  APA PsycInfo (Proquest, 1806 onwards)
  Public Health Database (Proquest, 1946 onwards)
  Psychology Database (Proquest, 1946 onwards)

Nursing & Allied Health Database (Proquest, 1946 onwards)
British Nursing Index (Proquest, 1994 onwards)

Publicly Available Content Database‎ (Proquest, no dates given)

- CENTRAL & CDSR (*The Cochrane Library*, issues 7 & 8 respectively)
- Stem Cell Evidence (Evidentia, 1950 onwards)
- Web of Science - All Databases except MEDLINE (Clarivate, 1900 onwards)

  *Ongoing Trial Databases:*
- ClinicalTrials.gov
- WHO International Clinical Trials Registry Platform (ICTRP)
- CENTRAL (*The Cochrane Library*, issue 7)

Searches were limited to English language only, and in MEDLINE and Embase will be combined with the NICE validated UK Geographic Search Filters (Ayiku 2017; Ayiku 2019) and in the Proquest and Web of Science databases with their respective UK location filters. (Searches in The Cochrane Library and Stem Cell Evidence are not able to be limited by location).

Search retrieved 11,574 references and 1,567 ongoing trials, which were reduced to 9,670 references and 1,020 ongoing trials once duplicates and (for ongoing trials) clearly non-UK studies had been removed in EndNote. These were then imported into Covidence for screening by Zareen Deplano on 28.8.24, where a further 45 duplicates were removed, leaving 10,645 references in total.

**SEARCH STRATEGIES**

**MEDLINE**

1.	exp Socioeconomic Factors/

2.	exp Health Inequities/

3.	Health Services Accessibility/

4.	"Social Determinants of Health"/

5.	exp Social Conditions/

6.	exp Social Deprivation/

7.	exp Social Environment/

8.	Sociodemographic Factors/

9.	Social Isolation/

10.	Social Marginalization/

11.	Social Vulnerability/

12.	((socio-economic* or socioeconomic* or sociodemograph* or socio-demograph* or social* or economic* or financ* or income* or wealth* or educat* or literacy or health or employ* or unemploy* or occupation* or job* or housing or environment* or geograph* or regional* or rural* or urban* or demograph*) adj3 (depriv* or inequalit* or inequit* or equit* or equalit* or status or position* or factor* or characteristic* or disparit* or variation* or barrier* or hardship* or determinant* or disadvantag* or difference* or class or classes or vulnerab* or insecur* or instabilit*)).tw,kf.

13.	((educat* or employ* or unemploy* or occupation* or job*) adj3 (status or disparit* or insecur*)).tw,kf.

14.	(poverty* or overcrowd* or over-crowd* or (depriv* adj2 (area* or region* or communit* or neighbo?rhood*)) or high-income* or middle-income* or low income* or low wage* or wealth difference* or wealth disparit* or unemployment or unemployed or underclass* or under-class* or jobless* or low education or social support or income support or household-income or household condition* or housing condition* or housing status or accessible housing or social condition or social conditions or poor household* or income tier* or ((geograph* or region*) adj2 (variation* or variabilit* or program* or boundar*)) or ((rural or urban) adj2 (access* or provid* or provision or communit* or area* or residen* or neighbo?rhood*)) or (food adj2 insecurit*) or car owner* or ((internet or broadband) adj2 (availab* or afford* or access* or provid* or provision))).tw,kf.

15.	((racial* or ethnic*) adj2 (inequali* or equali* or prejudi* or discriminat* or disadvantag* or disparit* or factor*)).tw,kf.

16.	((access* or provision or allocat* or referr* or wait* or list*) adj3 (inequalit* or inequit* or equit* or equalit* or status* or target* or threshold* or disparit* or variation* or barrier* or determinant* or disadvantag* or lower*)).tw,kf.

17.	(circle* adj2 (allocat* or polic* or based)).tw,kf.

18.	((social* or socio* or economic* or disadvanged or depriv* or poverty* or employ* or underprivileged or under-privileged or income* or racial or ethnic or minority) adj1 (group* or status or demograph* or factor* or position* or class*)).mp.

19.	or/1-18

20.	exp Stem Cell Transplantation/

21.	Bone Marrow Transplantation/

22.	Cell Transplantation/

23.	((stem cell* or marrow cell* or bone marrow or hematopoietic cell* or haematopoietic cell* or haematopoetic cell* or hematopoetic cell* or hemopoietic cell* or haemopoietic cell* or progenitor cell* or precursor cell* or peripheral blood) and (transplant* or graft* or engraft* or rescu* or allograft* or allo-graft* or pre-transplant* or pretransplant* or post-transplant* or posttransplant*)).tw,kf.

24.	(cell transplantation or stem cell* or bone marrow transplantation).jn.

25.	(((autologous or allogeneic or allogenic) adj5 transplant*) or cell* therap*).tw,kf.

26.	(allo-SCT or auto-SCT or ASCT or allo-BMT or auto-BMT or ABMT or PBPC or PBSCT or PSCT or BMT or SCT or HSCT or H-SCT).tw,kf.

27.	or/20-26

28.	exp Hematologic Neoplasms/

29.	exp Lymphoma/

30.	exp Leukemia/

31.	exp Neoplasms, Plasma Cell/

32.	exp Bone Marrow Diseases/

33.	((haematolog* or hematolog* or haemato-oncolog* or hemato-oncolog*) adj2 patients).tw,kf.

34.	(leukemi* or leukaemi* or lymphom* or myelodysplas* or myeloproliferat* or myelom* or plasm??ytom* or plasma cell dyscrasia* or POEMS).tw,kf.

35.	(lymphogranulomato* or histiocy* or granulom* or thrombocythemi* or thrombocythaemi* or polycythemi* or polycythaemi* or myelofibros* or AML or CLL or CML or ALCL or DLBCL or Hodgkin* or nonhodgkin* or B-NHL or waldenstrom* macroglobulinemia or reticulosis or reticulosarcom*).tw,kf.

36.	((burkitt* adj (lymph* or tumo?r)) or lymphosarcom* or brill-symmer* or sezary).tw,kf.

37.	((haematolog* or hematolog* or blood or red cell* or white cell* or lymph* or marrow or platelet*) adj3 (malignan* or oncolog* or cancer* or neoplasm* or carcinoma* or tumo?r*)).tw,kf.

38.	or/28-37

39.	19 and (27 or 38)

40.	exp Great Britain/

41.	(national health service* or nhs*).ti,ab,in.

42.	(english not ((published or publication* or translat* or written or language* or speak* or literature or citation*) adj5 english)).ti,ab.

43.	(gb or "g.b." or britain* or (british* not "british columbia") or uk or "u.k." or united kingdom* or (england* not "new england") or northern ireland* or northern irish* or scotland* or scottish* or ((wales or "south wales") not "new south wales") or welsh*).ti,ab,jw,in.

44.	(bath or "bath's" or ((birmingham not alabama*) or ("birmingham's" not alabama*) or bradford or "bradford's" or brighton or "brighton's" or bristol or "bristol's" or carlisle* or "carlisle's" or (cambridge not (massachusetts* or boston* or harvard*)) or ("cambridge's" not (massachusetts* or boston* or harvard*)) or (canterbury not zealand*) or ("canterbury's" not zealand*) or chelmsford or "chelmsford's" or chester or "chester's" or chichester or "chichester's" or coventry or "coventry's" or derby or "derby's" or (durham not (carolina* or nc)) or ("durham's" not (carolina* or nc)) or ely or "ely's" or exeter or "exeter's" or gloucester or "gloucester's" or hereford or "hereford's" or hull or "hull's" or lancaster or "lancaster's" or leeds* or leicester or "leicester's" or (lincoln not nebraska*) or ("lincoln's" not nebraska*) or (liverpool not (new south wales* or nsw)) or ("liverpool's" not (new south wales* or nsw)) or (((london not (ontario* or ont or toronto*)) or ("london's" not (ontario* or ont or toronto*)) or manchester or "manchester's" or (newcastle not (new south wales* or nsw)) or "newcastle's") not (new south wales* or nsw)) or norwich or "norwich's" or nottingham or "nottingham's" or oxford or "oxford's" or peterborough or "peterborough's" or plymouth or "plymouth's" or portsmouth or "portsmouth's" or preston or "preston's" or ripon or "ripon's" or salford or "salford's" or salisbury or "salisbury's" or sheffield or "sheffield's" or southampton or "southampton's" or "st albans" or stoke or "stoke's" or sunderland or "sunderland's" or truro or "truro's" or wakefield or "wakefield's" or wells or westminster or "westminster's" or winchester or "winchester's" or wolverhampton or "wolverhampton's" or (worcester not (massachusetts* or boston* or harvard*)) or ("worcester's" not (massachusetts* or boston* or harvard*)) or (york not ("new york*" or ny or ontario* or ont or toronto*)) or ("york's" not ("new york*" or ny or ontario* or ont or toronto*)))).ti,ab,in.

45.	(bangor or "bangor's" or cardiff or "cardiff's" or newport or "newport's" or "st asaph" or "st asaph's" or st davids or swansea or "swansea's").ti,ab,in.

46.	(aberdeen or "aberdeen's" or dundee or "dundee's" or edinburgh or "edinburgh's" or glasgow or "glasgow's" or inverness or (perth not australia*) or ("perth's" not australia*) or stirling or "stirling's").ti,ab,in.

47.	(armagh or "armagh's" or belfast or "belfast's" or lisburn or "lisburn's" or londonderry or "londonderry's" or derry or "derry's" or newry or "newry's").ti,ab,in.

48.	40 or 41 or 42 or 43 or 44 or 45 or 46 or 47

49.	(exp africa/ or exp americas/ or exp antarctic regions/ or exp arctic regions/ or exp asia/ or exp oceania/) not (exp great britain/ or europe/)

50.	48 not 49

51.	39 and 50

52.	(exp Animals/ or exp Animal Experimentation/ or exp Models, Animal/) not Humans/

53.	51 not 52

54.	limit 53 to english language

**Embase**

1.	socioeconomics/

2.	socioeconomic parameters/

3.	exp socioeconomic distribution/

4.	exp employment status/

5.	exp economic status/

6.	exp educational status/

7.	housing instability/

8.	exp social inequality/

9.	exp social equity/

10.	exp social status/

11.	exp social environment/

12.	sociodemographics/

13.	digital divide/

14.	poverty level/

15.	health care access/

16.	"social determinants of health"/

17.	social exclusion/

18.	social vulnerability/

19.	((socio-economic* or socioeconomic* or sociodemograph* or socio-demograph* or social* or economic* or financ* or income* or wealth* or educat* or literacy or health or employ* or unemploy* or occupation* or job* or housing or environment* or geograph* or regional* or rural* or urban* or demograph*) adj3 (depriv* or inequalit* or inequit* or equit* or equalit* or status or position* or factor* or characteristic* or disparit* or variation* or barrier* or hardship* or determinant* or disadvantag* or difference* or class or classes or vulnerab* or insecur* or instabilit*)).tw,kf.

20.	((educat* or employ* or unemploy* or occupation* or job*) adj3 (status or disparit* or insecur*)).tw,kf.

21.	(poverty* or overcrowd* or over-crowd* or (depriv* adj2 (area* or region* or communit* or neighbo?rhood*)) or high-income* or middle-income* or low income* or low wage* or wealth difference* or wealth disparit* or unemployment or unemployed or underclass* or under-class* or jobless* or low education or social support or income support or household-income or household condition* or housing condition* or housing status or accessible housing or social condition or social conditions or poor household* or income tier* or ((geograph* or region*) adj2 (variation* or variabilit* or program* or boundar*)) or ((rural or urban) adj2 (access* or provid* or provision or communit* or area* or residen* or neighbo?rhood*)) or (food adj2 insecurit*) or car owner* or ((internet or broadband) adj2 (availab* or afford* or access* or provid* or provision))).tw,kf.

22.	((racial* or ethnic*) adj2 (inequali* or equali* or prejudi* or discriminat* or disadvantag* or disparit* or factor*)).tw,kf.

23.	((access* or provision or allocat* or referr* or wait* or list*) adj2 (inequalit* or inequit* or equit* or equalit* or status* or target* or threshold* or disparit* or variation* or barrier* or determinant* or disadvantag* or lower*)).tw,kf.

24.	(circle* adj2 (allocat* or polic* or based)).tw,kf.

25.	((social* or socio* or economic* or disadvanged or depriv* or poverty* or employ* or underprivileged or under-privileged or income* or racial or ethnic or minority) adj1 (group* or status or demograph* or factor* or position* or class*)).mp.

26.	or/1-25

27.	stem cell transplantation/ or exp allogeneic stem cell transplantation/ or exp autologous stem cell transplantation/ or exp hematopoietic stem cell transplantation/ or exp peripheral blood stem cell transplantation/

28.	bone marrow transplantation/ or allogenic bone marrow transplantation/ or autologous bone marrow transplantation/

29.	((stem cell* or marrow cell* or bone marrow or hematopoietic cell* or haematopoietic cell* or haematopoetic cell* or hematopoetic cell* or hemopoietic cell* or haemopoietic cell* or progenitor cell* or precursor cell* or peripheral blood) and (transplant* or graft* or engraft* or rescu* or allograft* or allo-graft* or pre-transplant* or pretransplant* or post-transplant* or posttransplant*)).tw,kf.

30.	(cell transplantation or stem cell* or bone marrow transplantation).jn.

31.	(((autologous or allogeneic or allogenic) adj5 transplant*) or cell* therap*).tw,kf.

32.	(allo-SCT or auto-SCT or ASCT or allo-BMT or auto-BMT or ABMT or PBPC or PBSCT or PSCT or BMT or SCT or HSCT or H-SCT).tw,kf.

33.	or/27-32

34.	exp hematologic malignancy/

35.	exp myelodysplastic syndrome/

36.	POEMS syndrome/

37.	((haematolog* or hematolog* or haemato-oncolog* or hemato-oncolog*) adj2 patients).tw,kf.

38.	(leukemi* or leukaemi* or lymphom* or myelodysplas* or myeloproliferat* or myelom* or plasm??ytom* or plasma cell dyscrasia* or POEMS).tw,kf.

39.	(lymphogranulomato* or histiocy* or granulom* or thrombocythemi* or thrombocythaemi* or polycythemi* or polycythaemi* or myelofibros* or AML or CLL or CML or ALCL or DLBCL or Hodgkin* or nonhodgkin* or B-NHL or waldenstrom* macroglobulinemia or reticulosis or reticulosarcom*).tw,kf.

40.	((burkitt* adj (lymph* or tumo?r)) or lymphosarcom* or brill-symmer* or sezary).tw,kf.

41.	((haematolog* or hematolog* or blood or red cell* or white cell* or lymph* or marrow or platelet* or plasma cell*) adj3 (malignan* or oncolog* or cancer* or neoplasm* or carcinoma* or tumo?r*)).tw,kf.

42.	or/34-41

43.	26 and (33 or 42)

44.	united kingdom/

45.	(national health service* or nhs*).ti,ab,in,ad.

46.	(english not ((published or publication* or translat* or written or language* or speak* or literature or citation*) adj5 english)).ti,ab.

47.	(gb or "g.b." or britain* or (british* not "british columbia") or uk or "u.k." or united kingdom* or (england* not "new england") or northern ireland* or northern irish* or scotland* or scottish* or ((wales or "south wales") not "new south wales") or welsh*).ti,ab,jx,in,ad.

48.	(bath or "bath's" or ((birmingham not alabama*) or ("birmingham's" not alabama*) or bradford or "bradford's" or brighton or "brighton's" or bristol or "bristol's" or carlisle* or "carlisle's" or (cambridge not (massachusetts* or boston* or harvard*)) or ("cambridge's" not (massachusetts* or boston* or harvard*)) or (canterbury not zealand*) or ("canterbury's" not zealand*) or chelmsford or "chelmsford's" or chester or "chester's" or chichester or "chichester's" or coventry or "coventry's" or derby or "derby's" or (durham not (carolina* or nc)) or ("durham's" not (carolina* or nc)) or ely or "ely's" or exeter or "exeter's" or gloucester or "gloucester's" or hereford or "hereford's" or hull or "hull's" or lancaster or "lancaster's" or leeds* or leicester or "leicester's" or (lincoln not nebraska*) or ("lincoln's" not nebraska*) or (liverpool not (new south wales* or nsw)) or ("liverpool's" not (new south wales* or nsw)) or (((london not (ontario* or ont or toronto*)) or ("london's" not (ontario* or ont or toronto*)) or manchester or "manchester's" or (newcastle not (new south wales* or nsw)) or "newcastle's") not (new south wales* or nsw)) or norwich or "norwich's" or nottingham or "nottingham's" or oxford or "oxford's" or peterborough or "peterborough's" or plymouth or "plymouth's" or portsmouth or "portsmouth's" or preston or "preston's" or ripon or "ripon's" or salford or "salford's" or salisbury or "salisbury's" or sheffield or "sheffield's" or southampton or "southampton's" or "st albans" or stoke or "stoke's" or sunderland or "sunderland's" or truro or "truro's" or wakefield or "wakefield's" or wells or westminster or "westminster's" or winchester or "winchester's" or wolverhampton or "wolverhampton's" or (worcester not (massachusetts* or boston* or harvard*)) or ("worcester's" not (massachusetts* or boston* or harvard*)) or (york not ("new york*" or ny or ontario* or ont or toronto*)) or ("york's" not ("new york*" or ny or ontario* or ont or toronto*)))).ti,ab,in.

49.	(bangor or "bangor's" or cardiff or "cardiff's" or newport or "newport's" or "st asaph" or "st asaph's" or st davids or swansea or "swansea's").ti,ab,in,ad.

50.	(aberdeen or "aberdeen's" or dundee or "dundee's" or edinburgh or "edinburgh's" or glasgow or "glasgow's" or inverness or (perth not australia*) or ("perth's" not australia*) or stirling or "stirling's").ti,ab,in,ad.

51.	(armagh or "armagh's" or belfast or "belfast's" or lisburn or "lisburn's" or londonderry or "londonderry's" or derry or "derry's" or newry or "newry's").ti,ab,in,ad.

52.	or/44-51

53.	(exp "arctic and antarctic"/ or exp oceanic regions/ or exp western hemisphere/ or exp africa/ or exp asia/) not (united kingdom/ or europe/)

54.	52 not 53

55.	43 and 54

56.	(rat or rats or mouse or mice or swine or porcine or murine or sheep or lambs or pigs or piglets or rabbit or rabbits or cat or cats or dog or dogs or cattle or bovine or monkey or monkeys or trout or marmoset$1).ti. and animal experiment/

57.	animal experiment/ not (human experiment/ or human/)

58.	56 or 57

59.	55 not 58

60.	limit 59 to (english language and "remove medline records")

**PROQUEST DATABASES:**

#1 (subject("Stem Cells") OR noft((("stem cell" OR "stem cells" OR "bone marrow" OR "hematopoietic cell" OR "hematopoietic cells" OR "haematopoietic cell" OR "haematopoietic cells" OR "haematopoetic cell" OR "haematopoetic cells" OR "hematopoetic cell" OR "hematopoetic cells" OR "hemopoietic cell" OR "hemopoietic cells" OR "haemopoietic cell" OR "haemopoietic cells" OR "progenitor cell" OR "progenitor cells" OR "precursor cell" OR "precursor cells" OR "peripheral blood") AND (transplant* OR graft* OR engraft* OR rescu* OR allograft* OR allo-graft* OR pre-transplant* OR pretransplant* OR post-transplant* OR posttransplant*))) OR noft((((autologous OR allogeneic OR allogenic) NEAR/5 transplant*) OR "cell therapy" OR "cell therapies" OR "cellular therapy" OR "cellular therapies")) OR noft((allo-SCT OR auto-SCT OR ASCT OR allo-BMT OR auto-BMT OR ABMT OR PBPC OR PBSCT OR PSCT OR BMT OR SCT OR HSCT OR H-SCT)) OR subject(Leukemias) OR noft(((haematolog* OR hematolog* OR haemato-oncolog* OR hemato-oncolog*) NEAR/2 patients)) OR noft((leukemi* OR leukaemi* OR lymphom* OR myelodysplas* OR myeloproliferat* OR myelom* OR plasm??ytom* OR plasma cell dyscrasia* OR POEMS)) OR noft((lymphogranulomato* OR histiocy* OR granulom* OR thrombocythemi* OR thrombocythaemi* OR polycythemi* OR polycythaemi* OR myelofibros* OR AML OR CLL OR CML OR ALCL OR DLBCL OR Hodgkin* OR nonhodgkin* OR B-NHL OR waldenstrom* macroglobulinemia OR reticulosis OR reticulosarcom*)) OR noft(((burkitt* NEAR/1 (lymph* OR tumo?r)) OR lymphosarcom* OR brill-symmer* OR sezary)) OR noft(((haematolog* OR hematolog* OR blood OR "red cell" OR "red cells" OR "white cell" OR "white cells" OR lymph* OR marrow OR platelet*) NEAR/3 (malignan* OR oncolog* OR cancer* OR neoplasm* OR carcinoma* OR tumo?r*)))) AND lo.Exact("Great Britain" OR "Wales" OR "England" OR "United Kingdom" OR "Scotland" OR "Northern Ireland")

#2 ((social* OR economic* OR financ* OR income* OR wealth* OR educat* OR literacy OR health OR employ* OR unemploy* OR occupation* OR job* OR housing OR environment* OR geograph* OR regional* OR rural* OR urban* OR demograph*) AND (depriv* OR inequalit* OR inequit* OR equit* OR equalit* OR status OR position* OR factor* OR characteristic* OR disparit* OR variation* OR barrier* OR hardship* OR determinant* OR disadvantag* OR difference* OR class OR classes OR vulnerab* OR insecur* OR instabilit*))

#3 (socio-economic* OR socioeconomic* OR sociodemograph* OR socio-demograph* OR deprived OR deprivation*) OR ((educat* OR employ* OR unemploy* OR occupation* OR job*) NEAR/2 (status OR disparit* OR insecur*)) OR (poverty* OR overcrowd* OR over-crowd* OR (depriv* NEAR/2 (area* OR region* OR communit* OR neighbo?rhood*)) OR high-income* OR middle-income* OR low income* OR low wage* OR wealth difference* OR wealth disparit* OR unemployment OR unemployed OR underclass* OR under-class* OR jobless* OR low education OR social support OR income support OR household-income OR household condition* OR housing condition* OR housing status OR accessible housing OR social condition OR social conditions OR poor household* OR income tier*)

#4 ((geograph* OR region*) NEAR/2 (variation* OR variabilit* OR program* OR boundar*)) OR ((rural OR urban) NEAR/2 (access* OR provid* OR provision OR communit* OR area* OR residen* OR neighbo?rhood*)) OR (food NEAR/2 insecurit*) OR car owner* OR ((internet OR broadband) NEAR/2 (availab* OR afford* OR access* OR provid* OR provision)) OR ((racial* OR ethnic*) NEAR/2 (inequali* OR equali* OR prejudi* OR discriminat* OR disadvantag* OR disparit* OR factor*))

#5 ((access* OR provision OR allocat* OR referr* OR wait* OR list*) NEAR/5 (inequalit* OR inequit* OR equit* OR equalit* OR status* OR target* OR threshold* OR disparit* OR variation* OR barrier* OR determinant* OR disadvantag* OR lower*)) OR (circle* NEAR/2 (allocat* OR polic* OR based)) OR ((social* OR socio* OR economic* OR disadvanged OR depriv* OR poverty* OR employ* OR underprivileged OR under-privileged OR income* OR racial OR ethnic OR minority) NEAR/1 (group* OR status OR demograph* OR factor* OR position* OR class*))

#6 #1 AND (#2 OR #3 OR #4 OR #5)

**THE COCHRANE LIBRARY**

#1	MeSH descriptor: [Socioeconomic Factors] explode all trees

#2	MeSH descriptor: [Health Inequities] explode all trees

#3	MeSH descriptor: [Health Services Accessibility] this term only

#4	MeSH descriptor: [Social Determinants of Health] this term only

#5	MeSH descriptor: [Social Conditions] explode all trees

#6	MeSH descriptor: [Social Deprivation] explode all trees

#7	MeSH descriptor: [Social Environment] explode all trees

#8	MeSH descriptor: [Sociodemographic Factors] this term only

#9	MeSH descriptor: [Social Isolation] explode all trees

#10	MeSH descriptor: [Social Marginalization] this term only

#11	MeSH descriptor: [Social Vulnerability] this term only

#12	((socio-economic* or socioeconomic* or sociodemograph* or socio-demograph* or social* or economic* or financ* or income* or wealth* or educat* or literacy or health or employ* or unemploy* or occupation* or job* or housing or environment* or geograph* or regional* or rural* or urban* or demograph*) near/3 (depriv* or inequalit* or inequit* or equit* or equalit* or status or position* or factor* or characteristic* or disparit* or variation* or barrier* or hardship* or determinant* or disadvantag* or difference* or class or classes or vulnerab* or insecur* or instabilit*)):ti,ab

#13	((educat* or employ* or unemploy* or occupation* or job*) near/3 (status or disparit* or insecur*)):ti,ab

#14	(poverty* or overcrowd* or over-crowd* or (depriv* near/2 (area* or region* or communit* or neighbo?rhood*)) or "low income" or "low incomes" or "low wage" or "low wages" or "wealth difference" or (wealth next disparit*) or unemployment or unemployed or underclass* or under-class* or jobless* or "low education" "or social support" or "income support" or household-income* or "household condition" or "household conditions" or "housing condition" or "housing conditions" or "housing status" or "accessible housing" or "social condition" or "social conditions" or "poor households" or "income tier" or "income tiers" or ((geograph* or region*) near/2 (variation* or variabilit* or program* or boundar*)) or ((rural or urban) near/2 (access* or provid* or provision or communit* or area* or residen* or neighbo?rhood*)) or (food near/2 insecurit*) or "car owner" or "car owners" or ((internet or broadband) near/2 (availab* or afford* or access* or provid* or provision))):ti,ab

#15	((racial* or ethnic*) near/2 (inequali* or equali* or prejudi* or discriminat* or disadvantag* or disparit* or factor*)):ti,ab

#16	((access* or provision or allocat* or referr* or wait* or list*) near/2 (inequalit* or inequit* or equit* or equalit* or status* or target* or threshold* or disparit* or variation* or barrier* or determinant* or disadvantag* or lower*)):ti,ab

#17	(circle* near/2 (allocat* or polic* or based)):ti,ab

#18	((social* or socio* or economic* or disadvanged or depriv* or poverty* or employ* or underprivileged or under-privileged or income* or racial or ethnic or minority) near/1 (group* or status or demograph* or factor* or position* or class*)):ti,ab

#19	#1 or #2 or #3 or #4 or #5 or #6 or #7 or #8 or #9 or #10 or #11 or #12 or #13 or #14 or #15 or #16 or #17 or #18

#20	MeSH descriptor: [Stem Cell Transplantation] explode all trees

#21	MeSH descriptor: [Bone Marrow Transplantation] this term only

#22	MeSH descriptor: [Cell Transplantation] this term only

#23	(("stem cell" or "stem cells" or "marrow cell" or "marrow cells" or "bone marrow" or ((hematopoietic or haematopoietic or haematopoetic or hematopoetic or hemopoietic or haemopoietic or progenitor or precursor) next cell*) or "peripheral blood") and (transplant* or graft* or engraft* or rescu* or allograft* or allo-graft* or pre-transplant* or pretransplant* or post-transplant* or posttransplant*)):ti,ab

#24	((autologous or allogeneic or allogenic) near/5 transplant*):ti,ab

#25	(allo-SCT or auto-SCT or ASCT or allo-BMT or auto-BMT or ABMT or PBPC or PBSCT or PSCT or BMT or SCT or HSCT or H-SCT):ti,ab

#26	#20 or #21 or #22 or #23 or #24 or #25

#27	MeSH descriptor: [Hematologic Neoplasms] explode all trees

#28	MeSH descriptor: [Lymphoma] explode all trees

#29	MeSH descriptor: [Leukemia] explode all trees

#30	MeSH descriptor: [Neoplasms, Plasma Cell] explode all trees

#31	MeSH descriptor: [Bone Marrow Diseases] explode all trees

#32	((haematolog* or hematolog* or haemato-oncolog* or hemato-oncolog*) near/2 patients):ti,ab

#33	(leukemi* or leukaemi* or lymphom* or myelodysplas* or myeloproliferat* or myelom* or plasm??ytom* or plasma cell dyscrasia* or POEMS):ti,ab

#34	(lymphogranulomato* or histiocy* or granulom* or thrombocythemi* or thrombocythaemi* or polycythemi* or polycythaemi* or myelofibros* or AML or CLL or CML or ALCL or DLBCL or Hodgkin* or nonhodgkin* or B-NHL or waldenstrom* macroglobulinemia or reticulosis or reticulosarcom*):ti,ab

#35	((burkitt* next (lymph* or tumo?r)) or lymphosarcom* or brill-symmer* or sezary):ti,ab

#36	((haematolog* or hematolog* or blood or red cell* or white cell* or lymph* or marrow or platelet*) near/3 (malignan* or oncolog* or cancer* or neoplasm* or carcinoma* or tumo?r*)):ti,ab

#37	#27 or #28 or #29 or #30 or #31 or #32 or #33 or #34 or #35 or #36

#38	#19 and (#26 or #37)

**STEM CELL EVIDENCE**

socio-economic OR socioeconomic OR sociodemographic OR socio-demographic OR social OR economic OR financial OR income OR wealth OR education OR literacy OR employment OR unemployment OR unemployed OR occupation OR job OR housing OR household OR environment OR geographical OR rural OR urban OR sociodemographic OR socio-demographic OR demographic OR deprivation OR deprived OR inequality OR inequity OR disparity OR disparities OR barrier OR disadvantage OR poverty OR insecurity

**WEB OF SCIENCE**

1. TS=(("stem cell" OR "stem cells" OR "bone marrow" OR "hematopoietic cell" OR "hematopoietic cells" OR "haematopoietic cell" OR "haematopoietic cells" OR "haematopoetic cell" OR "haematopoetic cells" OR "hematopoetic cell" OR "hematopoetic cells" OR "hemopoietic cell" OR "hemopoietic cells" OR "haemopoietic cell" OR "haemopoietic cells" OR "progenitor cell" OR "progenitor cells" OR "precursor cell" OR "precursor cells" OR "peripheral blood" OR autologous OR allogeneic OR allogenic) NEAR/5 (transplant* OR graft* OR engraft* OR rescu* OR allograft* OR allo-graft* OR pre-transplant* OR pretransplant* OR post-transplant* OR posttransplant*)) OR TI=(allo-SCT OR auto-SCT OR ASCT OR allo-BMT OR auto-BMT OR ABMT OR PBPC OR PBSCT OR PSCT OR BMT OR SCT OR HSCT OR H-SCT) OR AB=(allo-SCT OR auto-SCT OR ASCT OR allo-BMT OR auto-BMT OR ABMT OR PBPC OR PBSCT OR PSCT OR BMT OR SCT OR HSCT OR H-SCT)

2. TS=((haemat-oncolog* OR hemat-oncolog* OR haemato-oncolog* OR hemato-oncolog*) NEAR/1 patients) OR TI=(leukemi* OR leukaemi* OR lymphom* OR myelodysplas* OR myeloproliferat* OR myelom* OR plasm??ytom* OR "plasma cell dyscrasia" OR POEMS OR lymphogranulomato* OR histiocy* OR granulom* OR thrombocythemi* OR thrombocythaemi* OR polycythemi* OR polycythaemi* OR myelofibros* OR AML OR CLL OR CML OR ALCL OR DLBCL OR Hodgkin* OR nonhodgkin* OR non-Hodgkin* OR B-NHL OR waldenstrom* macroglobulinemia OR reticulosis OR reticulosarcom* OR lymphosarcom* OR brill-symmer* OR sezary) OR TI=(burkitt* NEAR/1 (lymph* OR tumo?r)) OR TI=((haematolog* OR hematolog* OR blood OR "red cell" OR "red cells" OR "white cell" OR "white cells" OR lymph* OR marrow OR platelet*) NEAR/3 (malignan* OR oncolog* OR cancer* OR neoplasm* OR carcinoma* OR tumo?r*)) OR AB=((haematolog* OR hematolog* OR blood OR "red cell" OR "red cells" OR "white cell" OR "white cells" OR lymph* OR marrow OR platelet*) NEAR/3 (malignan* OR oncolog* OR cancer* OR neoplasm* OR carcinoma* OR tumo?r*))

3. 1 OR 2

4. TS=((socio-economic* OR socioeconomic* OR sociodemograph* OR socio-demograph* OR social* OR economic* OR financ* OR income* OR wealth* OR educat* OR literacy OR health OR employ* OR unemploy* OR occupation* OR job* OR housing OR environment* OR geograph* OR regional* OR rural* OR urban* OR demograph*) NEAR/3 (depriv* OR inequalit* OR inequit* OR equit* OR equalit* OR status OR position* OR factor* OR characteristic* OR disparit* OR variation* OR barrier* OR hardship* OR determinant* OR disadvantag* OR difference* OR class OR classes OR vulnerab* OR insecur* OR instabilit*))

5. TI=(socio-economic* OR socioeconomic* OR sociodemograph* OR socio-demograph* OR "social inequality" OR "social inequalities" OR "social deprivation")

6. TS=((educat* OR employ* OR unemploy* OR occupation* OR job*) NEAR/3 (status OR disparit* OR insecur*))

7. TI=(poverty* OR overcrowd* OR over-crowd* OR (depriv* NEAR/2 (area* OR region* OR communit* OR neighbo?rhood*)) OR "low income" OR "low incomes" OR "low wage" OR "low wages" OR "wealth difference" OR "wealth disparity" OR "wealth disparities" OR unemployment OR unemployed OR underclass* OR under-class* OR jobless* OR "low education" OR "financial support" OR "financial assistance" OR "income support" OR "household income" OR "household condition" OR "household conditions" OR "housing condition" OR "housing conditions" OR "housing status" OR accessible housing" OR "social condition" OR "social conditions" OR "poor households" OR "income tier" OR "income tiers" OR "car owner" OR "car owners") OR AB=(poverty* OR overcrowd* OR over-crowd* OR (depriv* NEAR/2 (area* OR region* OR communit* OR neighbo?rhood*)) OR "low income" OR "low incomes" OR "low wage" OR "low wages" OR "wealth difference" OR "wealth disparity" OR "wealth disparities" OR unemployment OR unemployed OR underclass* OR under-class* OR jobless* OR "low education" OR "financial support" OR "financial assistance" OR "income support" OR "household income" OR "household condition" OR "household conditions" OR "housing condition" OR "housing conditions" OR "housing status" OR accessible housing" OR "social condition" OR "social conditions" OR "poor households" OR "income tier" OR "income tiers" OR "car owner" OR "car owners")

8. TS=((geograph* OR region*) NEAR/2 (variation* OR variabilit* OR program* OR boundar*)) OR TS=((rural OR urban) NEAR/2 (access* OR provid* OR provision OR communit* OR area* OR residen* OR neighbo?rhood*)) OR TS=(food NEAR/2 insecurit*)

9. TS=((internet OR broadband) NEAR/2 (availab* OR afford* OR access* OR provid* OR provision)) OR TS=((racial* OR ethnic*) NEAR/2 (inequali* OR equali* OR prejudi* OR discriminat* OR disadvantag* OR disparit* OR factor*))

10. TS=((access* OR provision OR allocat* OR referr* OR wait* OR list*) NEAR/2 (inequalit* OR inequit* OR equit* OR equalit* OR status* OR target* OR threshold* OR disparit* OR variation* OR barrier* OR determinant* OR disadvantag* OR lower*)) OR TS=(circle* NEAR/2 (allocat* OR polic* OR based)) OR TS=((social* OR socio* OR economic* OR disadvanged OR depriv* OR poverty* OR employ* OR underprivileged OR under-privileged OR income* OR racial OR ethnic OR minority) NEAR/1 (group* OR status OR demograph* OR factor* OR position* OR class*))

11. #4 OR #5 OR #6 OR #7 OR #8 OR #9 OR #10

12. #11 AND #3

13. #11 AND #3 and ENGLAND or WALES or NORTH IRELAND or SCOTLAND (Countries/Regions)

**ClinicalTrials.gov**

(Condition: hematological malignancy OR haematological malignancy OR hemato-oncology OR leukemia OR leukaemia OR lymphoma OR myelodysplastic OR myelodysplasia OR myeloproliferative OR myeloma OR hodgkin OR non-hodgkin OR plasmacytoma OR "plasma cell dyscrasia" OR POEMS

AND

Other Terms: socio-economic OR socioeconomic OR sociodemographic OR socio-demographic OR social OR economic OR financial OR income OR wealth OR education OR literacy OR employment OR unemployment OR unemployed OR occupation OR job OR housing OR household OR environment OR geographical OR rural OR urban OR sociodemographic OR socio-demographic OR demographic OR deprivation OR deprived OR inequality OR inequity OR disparity OR disparities OR barrier OR disadvantage OR poverty OR insecurity

AND

Location: United Kingdom)

OR

(Intervention: stem cell transplant OR stem cells OR bone marrow transplant OR hematopoietic cells OR haematopoietic cells OR haematopoetic cells OR hematopoetic cells OR hemopoietic cells OR haemopoietic cells OR progenitor cells OR precursor cells OR peripheral blood

AND

Other Terms: socio-economic OR socioeconomic OR sociodemographic OR socio-demographic OR social OR economic OR financial OR income OR wealth OR education OR literacy OR employment OR unemployment OR unemployed OR occupation OR job OR housing OR household OR environment OR geographical OR rural OR urban OR sociodemographic OR socio-demographic OR demographic OR deprivation OR deprived OR inequality OR inequity OR disparity OR disparities OR barrier OR disadvantage OR poverty OR insecurity

AND

Location: United Kingdom)

**WHO ICTRP**

(hematological malignancy OR haematological malignancy OR hemato-oncology OR leukemia OR leukaemia OR lymphoma OR myelodysplastic OR myelodysplasia OR myeloproliferative OR myeloma OR hodgkin OR non-hodgkin OR plasmacytoma OR "plasma cell dyscrasia" OR POEMS) AND (socio-economic OR socioeconomic OR sociodemographic OR socio-demographic OR social OR economic OR financial OR income OR wealth OR education OR literacy OR employment OR unemployment OR unemployed OR occupation OR job OR housing OR household OR environment OR geographical OR rural OR urban OR sociodemographic OR socio-demographic OR demographic OR deprivation OR deprived OR inequality OR inequity OR disparity OR disparities OR barrier OR disadvantage OR poverty OR insecurity)

OR

(stem cell transplant OR stem cells OR bone marrow transplant OR hematopoietic cells OR haematopoietic cells OR haematopoetic cells OR hematopoetic cells OR hemopoietic cells OR haemopoietic cells OR progenitor cells OR precursor cells OR peripheral blood) AND (socio-economic OR socioeconomic OR sociodemographic OR socio-demographic OR social OR economic OR financial OR income OR wealth OR education OR literacy OR employment OR unemployment OR unemployed OR occupation OR job OR housing OR household OR environment OR geographical OR rural OR urban OR sociodemographic OR socio-demographic OR demographic OR deprivation OR deprived OR inequality OR inequity OR disparity OR disparities OR barrier OR disadvantage OR poverty OR insecurity)
