## Supplementary File 3 for "The role of socioeconomic deprivation on inequities in care outcomes of haematological malignancies, treated with stem cell transplant in the UK: A systematic Review Protocol"

Supplementary File 4. Data extraction form

| **General information** | Data extractor |
| --- | --- |
|  | Date form completed |
|  | Study ID |
|  | First Author |
|  | Citation |
|  | Year of publication |
|  | Geographical location |
|  | Funding-conflict of interest |
|  | Information regarding ethical approval |
| **Methods** |  |
| **Study Eligibility and characteristics** | Title |
|  | Study type |
|  | Study design |
|  | Data Source |
|  | Study aim |
|  | Start and end date of study |
|  | Total study duration |
|  | Method of participant recruitment |
|  | Statistical methods and appropriateness of methods |
| **Population characteristics** | Selection criteria |
|  | Study size (number of patients) |
|  | Description of population |
|  | Clinical indication for SCT |
|  | Year of diagnosis |
|  | Genetics risk (ELN Standard, intermediate, adverse risk, grade  low grade/high grade lymphoma, myeloma ( International Staging System (ISS): stage I, II, III |
|  | Performance status ECOG reported (Y/N) |
|  | Age summary statistic (range/median and SD) |
|  | Sex or gender summary statistics (per cent women/female) |
|  | Ethnicity |
|  | Disease status (Complete remission/Partial remission/relapse) |
|  | Type of treatment received (intensive chemotherapy/conditioning or non-intensive chemo) |
|  | Geographical distance from TC reported (Y/N) |
|  | Treatment at local hospital vs tertiary transplant centre or academic institution |
|  | Co-morbidities (obesity, type 2 diabetes, lung disease, and  cardiovascular and cerebrovascular diseases) reported |
|  | Measurement of CI (Charlson or Hematopoietic cell transplantation-specific comorbidity index (HCTCI) scoring systems |
|  | Follow-­up duration |
| **Exposure and outcome** | Exposure definition: socio-economic deprivation: 1) individual-­level or household-­level socioeconomic factor such as employment, education, income, housing, poverty/low wage, social isolation and car ownership.  2) Aggregate area-­level measures of SES (such as the Index of Multiple Deprivation, Carstairs, Townsend index) |
|  | Measurement of assessment of socioeconomic deprivation (individual or neighbourhood level) |
|  | 2) Measure of socioeconomic deprivation Aggregate area-­level measures of SES (such as the Index of Multiple Deprivation, Townsend, Car stairs) |
|  | Type of deprivation (material or relative) |
|  | Outcome definition |
|  | Outcome measurement (number/%) |
| **Effect size and uncertainty** | Effect size measure (e.g., relative risk, OR, incidence rate ratio, HR) |
|  | Effect size |
|  | CI and level |
|  | Interval uncertainty type and value |
|  | Sample size (total, exposed, unexposed) |
|  | No. of events (total, among exposed, among unexposed) |
|  | Whether main or subgroup analysis |
|  | Description of subgroup analysis |
| **Main Findings** |  |
| **Risk of bias (good, fair and poor )** |  |
| NOS, Newcastle-­Ottawa Scale |  |
